# A rapid systematic review of measures to protect older people in long term care facilities from COVID-19

**DOI:** 10.1101/2020.10.29.20222182

**Authors:** Kate Frazer, Lachlan Mitchell, Diarmuid Stokes, Ella Lacey, Eibhlin Crowley, Cecily Kelleher

**Affiliations:** School of Nursing, Midwifery and Health Systems, University College Dublin, Belfield, Dublin 4, Ireland; National Nutrition Surveillance Centre, University College Dublin, Belfield, Dublin 4, Ireland; School of Public Health, Physiotherapy and Sport Science, University College Dublin, Belfield, Dublin 4, Ireland; Health Sciences Library, University College Dublin, Belfield, Dublin 4, Ireland; School of Medicine, University College Dublin, Belfield, Dublin 4, Ireland; Office for Health Affairs, College of Health and Agricultural Science, University College Dublin, Belfield, Dublin 4, Ireland

**Author notes:** Equal contribution (Kate Frazer, Lachlan Mitchell).

**Keywords:** Coronavirus, COVID-19, nursing homes, transmission

## Abstract

The global COVID-19 pandemic produced large-scale health and economic complications. Older people and those with comorbidities are particularly vulnerable to this virus, with nursing homes and long term care facilities experiencing significant morbidity and mortality associated with COVID-19 outbreaks. The aim of this rapid systematic review was to investigate measures implemented in long term care facilities to reduce transmission of COVID-19 and their effect on morbidity and mortality of residents, staff, and visitors. Databases (including MedRXiv pre-published repository) were systematically searched to identify studies reporting assessment of interventions to reduce transmission of COVID-19 in nursing homes among residents, staff, or visitors. Outcome measures include facility characteristics, morbidity data, case fatalities, and transmission rates. Due to study quality and heterogeneity, no meta-analysis was conducted. The search yielded 1414 articles, with 38 studies included. Reported interventions include mass testing, use of personal protective equipment, symptom screening, visitor restrictions, hand hygiene and droplet/contact precautions, and resident cohorting. Prevalence rates ranged from 1.2-85.4% in residents and 0.6-62.6% in staff. Mortality rates ranged from 5.3-55.3% in residents. Novel evidence in this review details the impact of facility size, availability of staff and practices of operating between multiple facilities, and for-profit status of facilities as factors contributing to the size and number of COVID-19 outbreaks. No causative relationships can be determined; however, this review provides evidence of interventions that reduce transmission of COVID-19 in long term care facilities.

## Introduction

Severe Acute Respiratory Syndrome Coronavirus-2 (SARS-CoV-2) is a novel virus, first identified in China in 2019, resulting in the current global pandemic in 2020.^1^ The ensuing disease associated with infection from SARS-CoV-2, termed COVID-19, has produced large-scale public health and worldwide economic effects.^2^

The virus spreads between people through close contact and droplet transmission (coughs and sneezes). While most infected people will experience mild flu□like symptoms, others may become seriously ill and die.^3^ At-risk groups include older people and those with underlying medical conditions, while men appear to have more susceptibility than women. Symptom severity varies; several individuals remain asymptomatic, others experience fever, cough, sore throat, general weakness, and fatigue, while more severe respiratory illnesses and infections may result, which can be fatal.^4,5^ Deterioration in clinical presentations can occur rapidly, leading to poorer health outcomes. Anosmia and ageusia are reported in evidence from South Korea, China, and Italy in patients with confirmed SARS-CoV-2 infection, in some cases in the absence of other symptoms.^6^

The World Health Organization (WHO) declared the COVID-19 outbreak constituted a Public Health Emergency of International Concern (PHEIC) on January 30, 2020.^5^ Two primary goals of action were 1) to accelerate innovative research to help contain the spread and facilitate care for all affected, and 2) to support research priorities globally the learning from the pandemic response for preparedness. Globally, up to October 5, 2020, there are 35 247 104 cases of COVID-19 (following the applied case definitions and testing strategies in the affected countries) including 1 038 069 deaths.^7^ Within Europe, over 5 431 510 cases are reported, with 226 869 deaths^7^ Presently there is no vaccine; therefore, preventing and limiting transmission is advocated. International and national evidence mandates physical distancing, regular hand hygiene and cough etiquette, and limiting touching eyes, nose or mouth; in addition to regular cleaning of surfaces.^8^

As noted older people are an at-risk group for COVID-19, and throughout the pandemic, the impact on this population has resulted in increased mortality, specifically those living in long term care facilities (LTCF) where a high proportion of outbreaks with increased rates of morbidity and case fatality in residents are recorded.^9^ In several EU/EEA countries, LTCF deaths among residents, associated with COVID-19, account for 37% to 66% of all COVID-19-related fatalities.^9^ The specific rationale for their increased susceptibility is less clear. The United Nations (UN) (2020) acknowledge that COVID-19 exposes the inequalities in society and the failures expressed in the 2030 Agenda for Sustainable Development. The UN report the disproportionate fatality rates in those aged over 80 years as five times the global average^10^ and suggest a need for a more *inclusive, equitable and age-friendly society, anchored in human rights* (p.16).^11^

The aim of this rapid review of the literature was to assess the extent to which measures implemented in LTCF reduced transmission of COVID-19 (SARS-CoV-2) among residents, staff, and visitors, and the effect of these measures on morbidity and mortality outcomes.

## Methods

The protocol is registered on PROSPERO (CRD42020191569)^12^ and reporting follows PRISMA guidelines.^13^

### Search strategy

Search strategies comprised search terms both for keywords and controlled-vocabulary search terms MESH and EMTREE (see Supplementary Table 1 for full search terms). EMBASE (via OVID), PubMed (via OVID), Cumulative Index to Nursing and Allied Health Literature (CINAHL), Cochrane Database and Repository, and MedRXiv pre-published databases were searched. No time limits were imposed, and databases were searched up to July 27, 2020. Reference lists of included evidence were checked for further articles.

### Eligibility criteria

All study designs (experimental, observational, and qualitative) are included, and no exclusions placed on language. Included studies report an assessment of measures to reduce transmission of COVID-19 (including SARS or MERS) in residents, employees, or visitors of LTCF. To provide as comprehensive a review of the evidence we included any intervention implemented to reduce the transmission of COVID-19 in long-term residential care facilities, including facility measures, social distancing, use of personal protective equipment, and hand hygiene.

### Primary outcome measures

Primary outcome measures are morbidity data, case fatality rates, reductions in reported transmission rates, and facility characteristics associated with COVID-19 incidence.

### Selection of studies and data extraction

Two authors developed search strings (DS & KF); all database searches were completed by one author (DS) (Supplementary Table 1). Following de-duplication, references were uploaded into Covidence management platform (LM), and two authors independently screened all titles and abstracts (LM & KF). Full texts of all potentially eligible studies were independently reviewed by two authors (LM & KF). Disagreements were resolved by discussion with a third author (CK). Data from included studies were independently extracted in duplicate (LM & KF). A data extraction form was developed and modified from documents used previously by authors (KF & CK). Extracted data included study characteristics (title, lead author, year of publication, country, study setting, study design), description of the intervention, number and characteristics of participants, outcomes, duration of follow-up, sources of funding, peer review status). Study design (required for review of quality) was independently assessed by two authors (LM & KF), with disagreements resolved by a third author (CK).

### Assessment of Quality

Two review authors (LM & EL) independently assessed the quality of included studies using Mixed Methods Assessment Tool (MMAT),^14^ with disagreements resolved by a third author (KF) and discussed with the lead author (CK) (Supplementary Table 2). The MMAT is used widely and considered a valid indicator of methodological quality using instruments for non-randomised and descriptive studies.

### Data synthesis

Meta-analysis was not possible due to heterogeneity in study designs, participants, outcomes, and nature of the interventions and no attempt was made to transform statistical data. The SWiM criteria^15^ guide a narrative summary, with data presented in tabular format and subgroup reporting of population groups.

## Results

We identified 1414 articles, and 131 full-text articles were selected for review. After an evaluation against our inclusion criteria, 38 studies (40 papers) are included in this systematic review (Figure 1).

**Figure 1.**
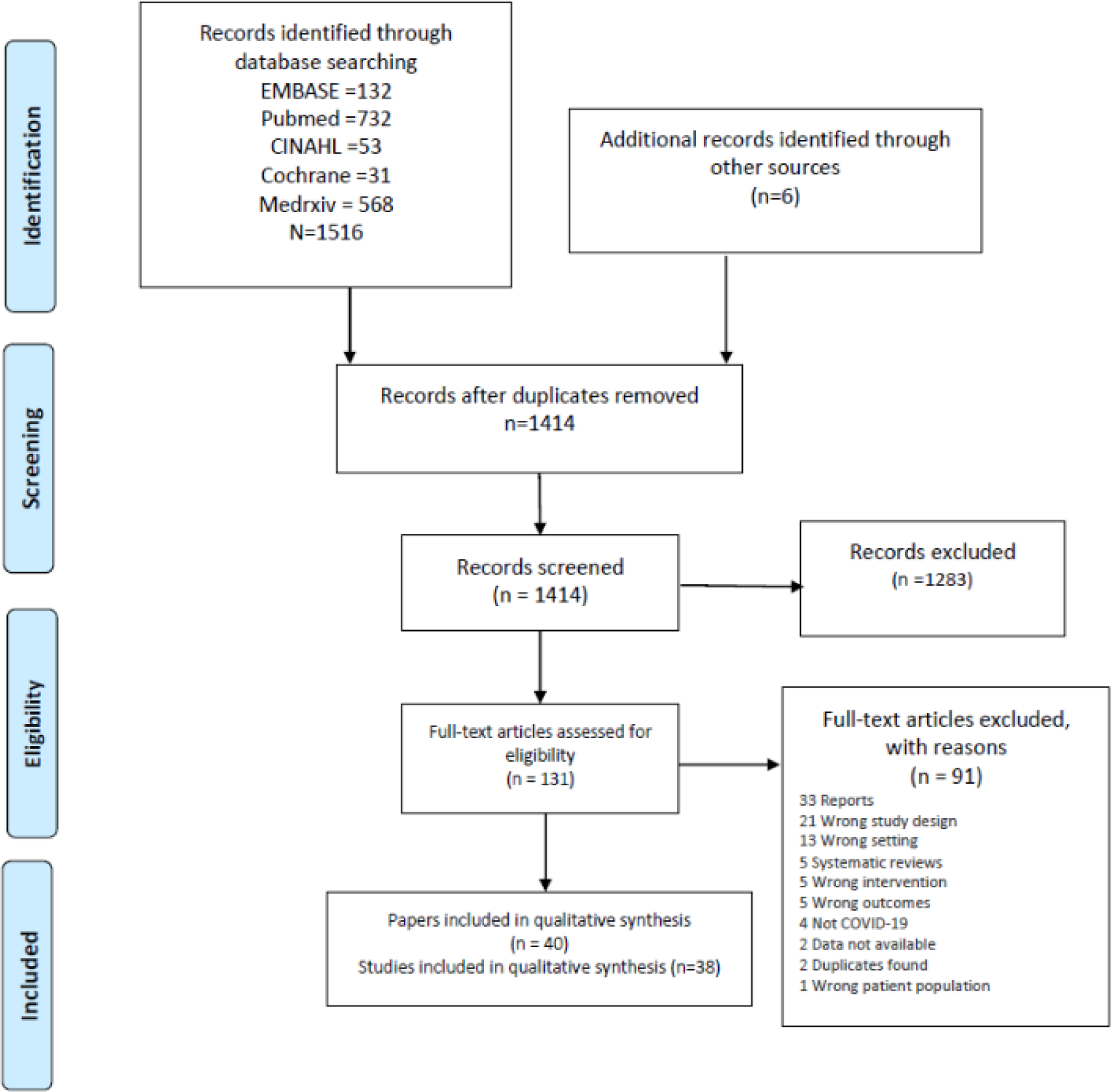
PRISMA Flowchart.

### Study characteristics

Geographically we report evidence from eleven countries, the majority (20 studies) are from USA^16-35^ and UK.^36-40^ We report evidence from Canada,^41-43^ France,^44,45^ Hong Kong,^46,47^ Belgium,^48^ Germany,^49^ Ireland,^50^ Japan,^51^ Korea,^52^ and Spain^53^ (Table 1).

**Table 1.**
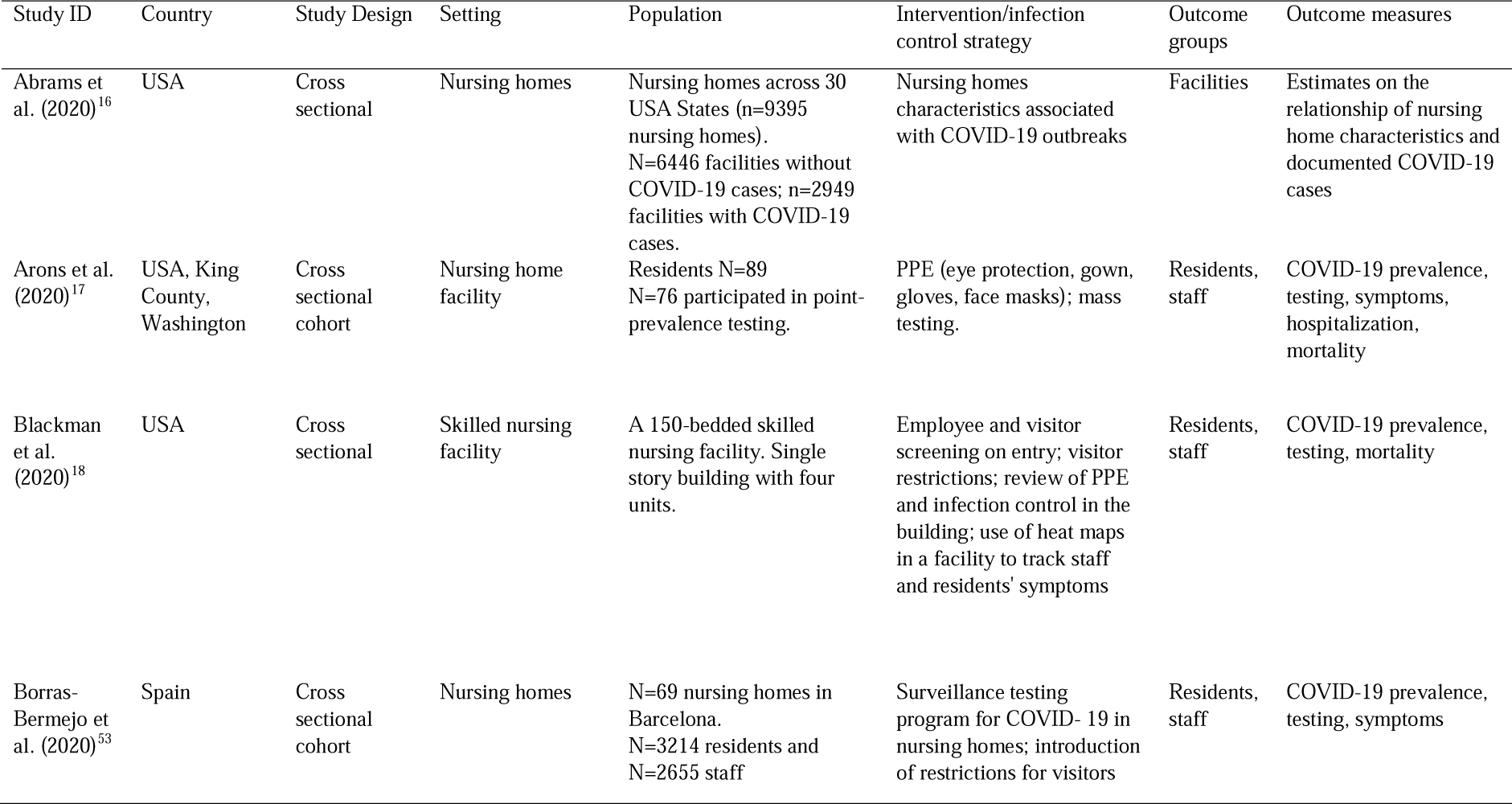
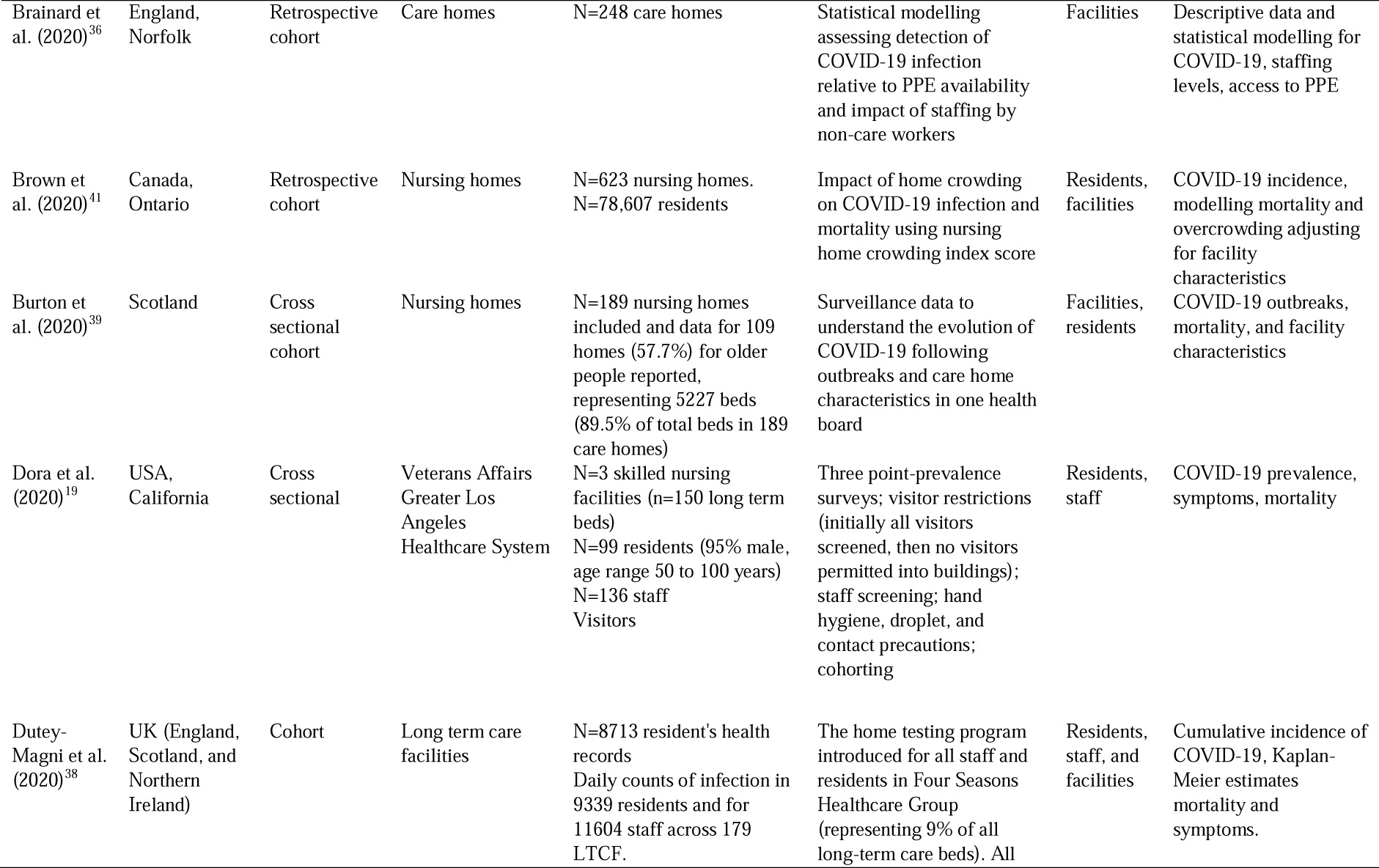
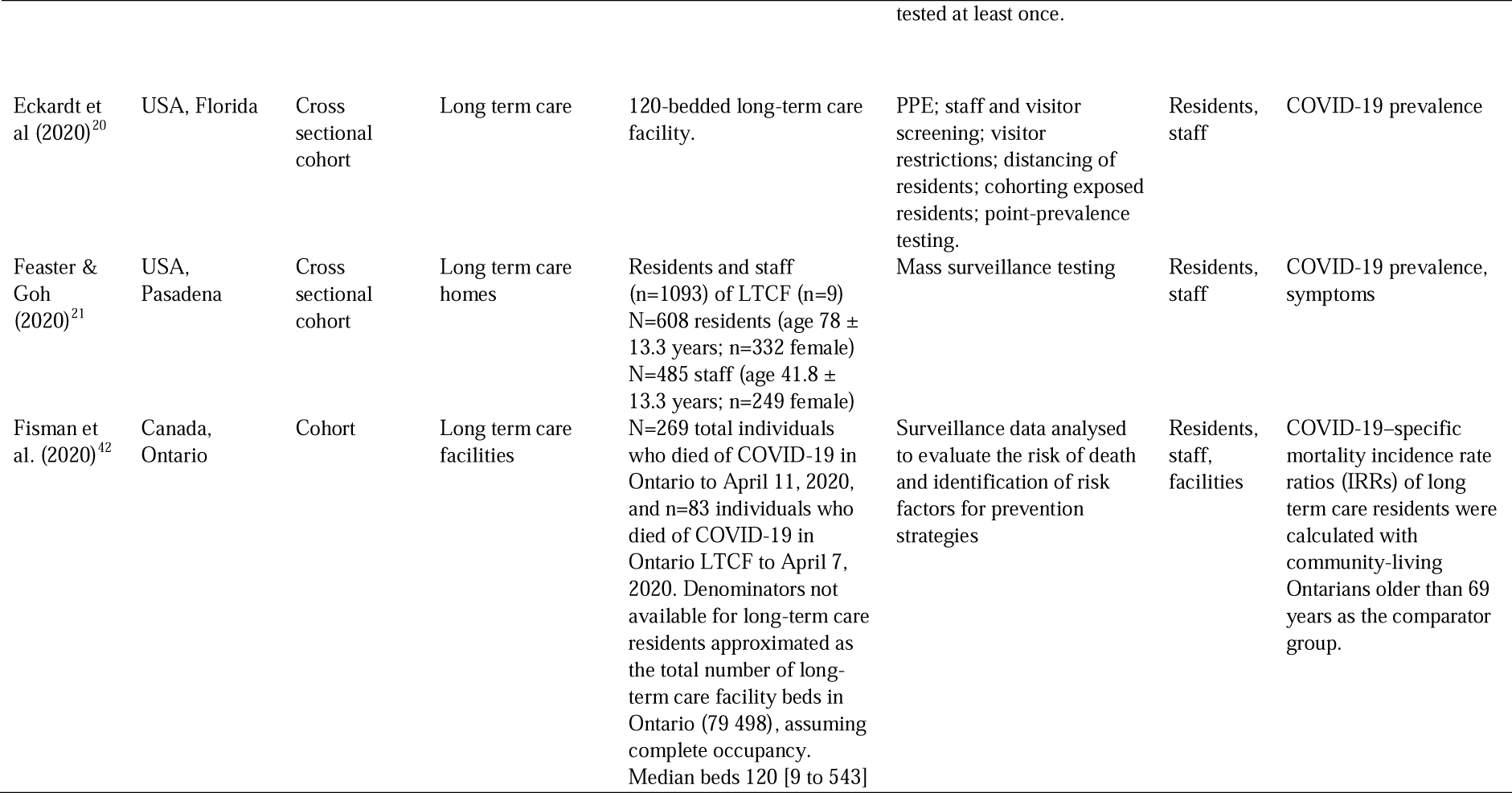
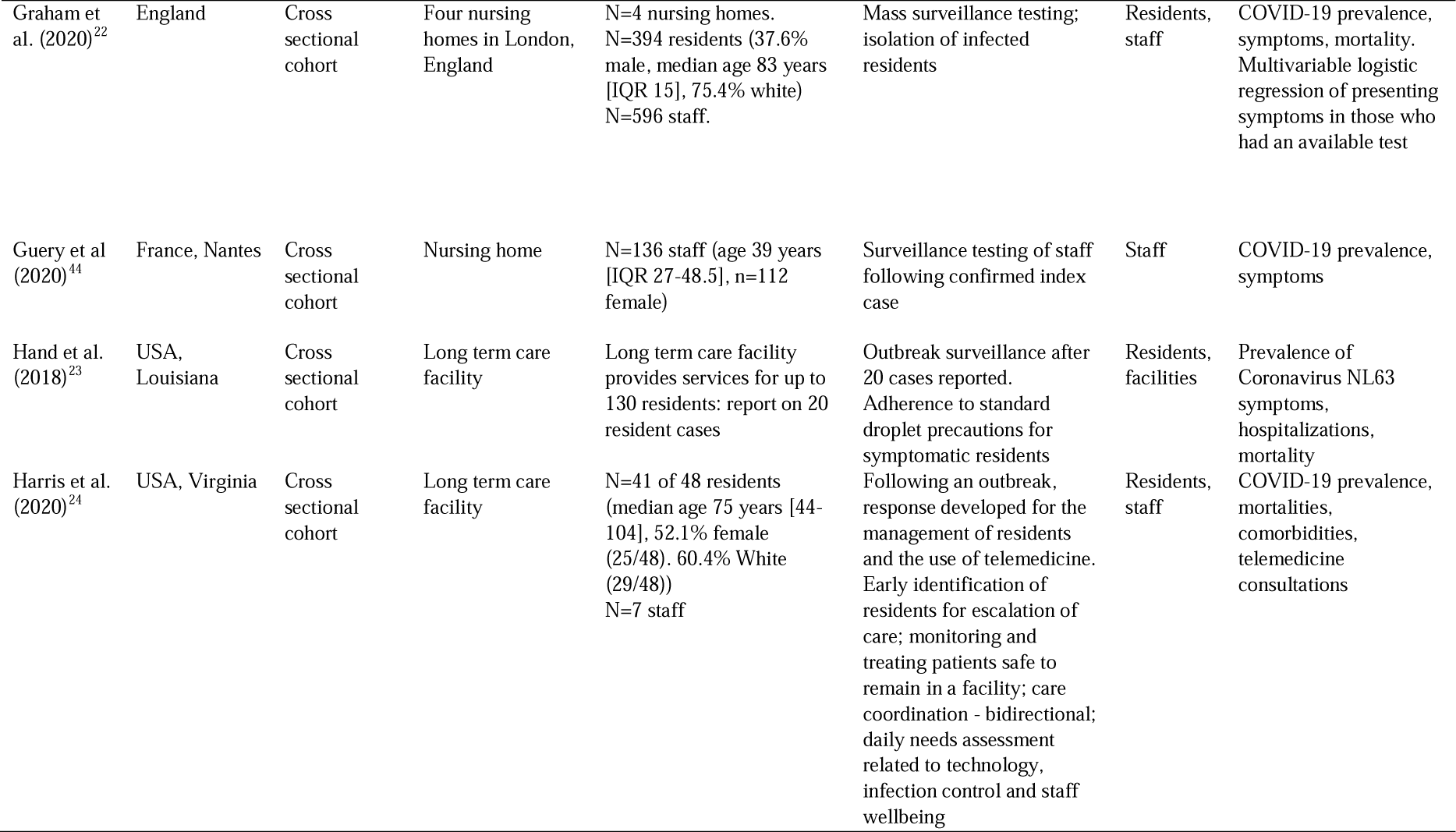
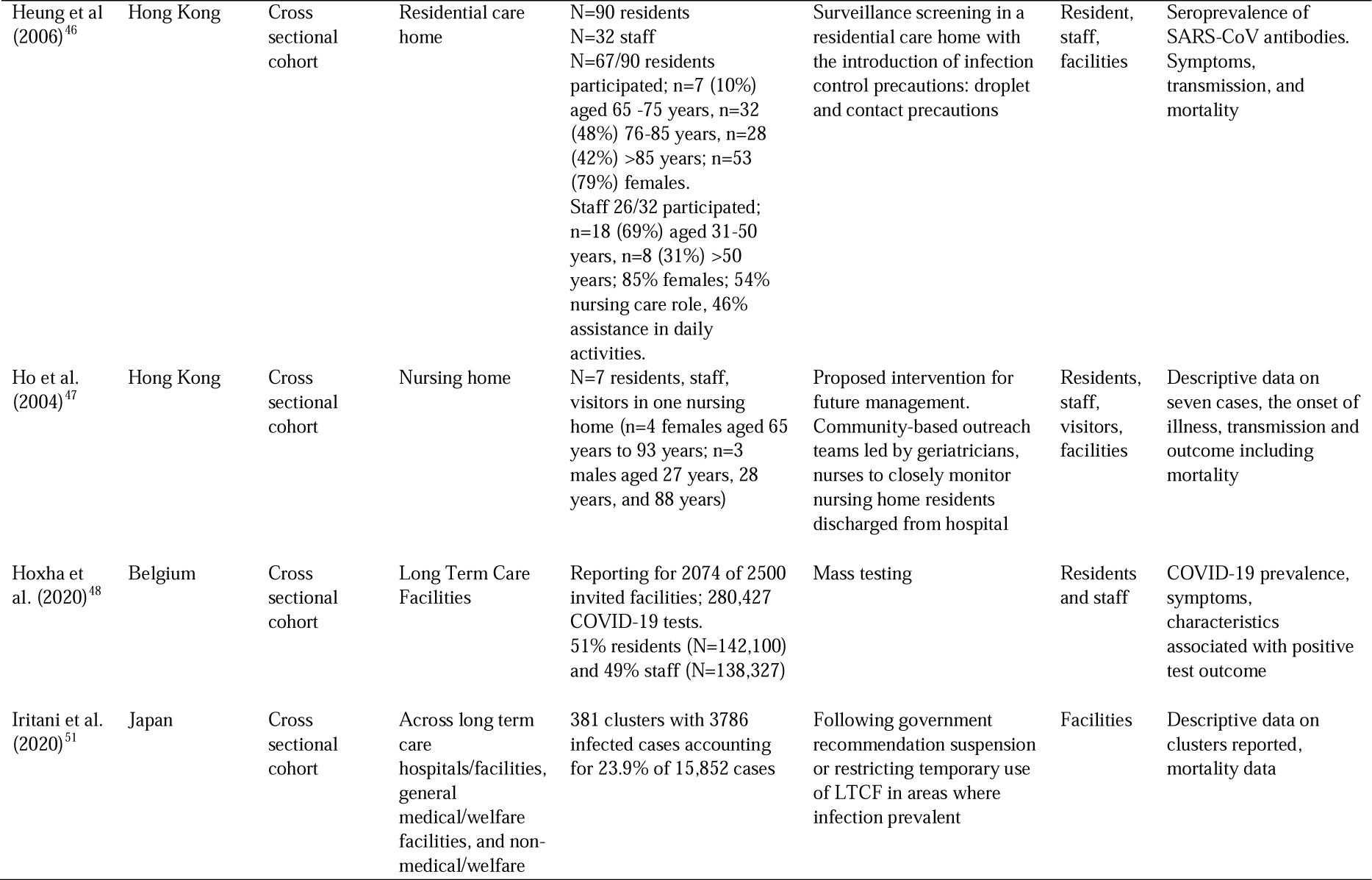
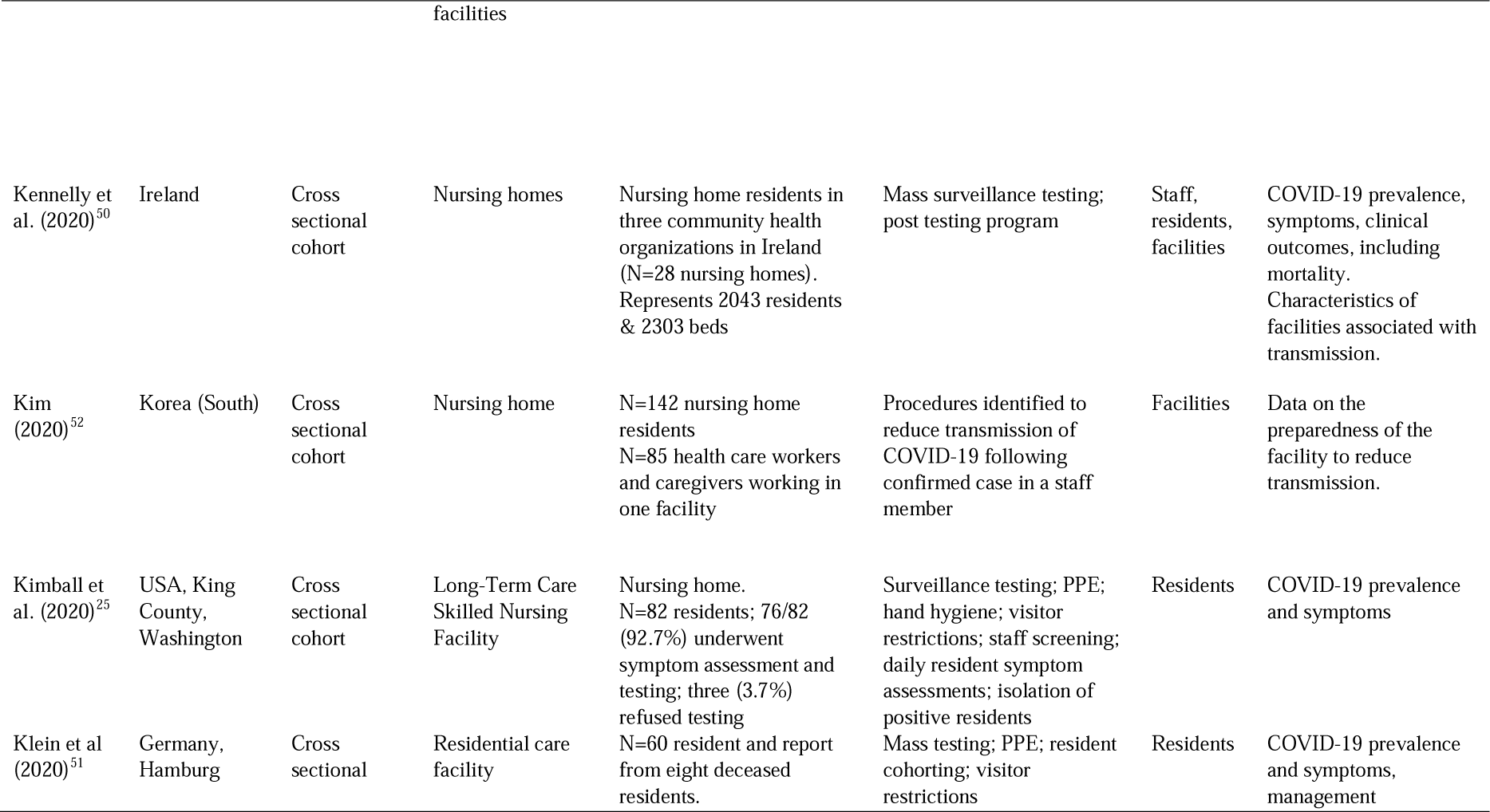
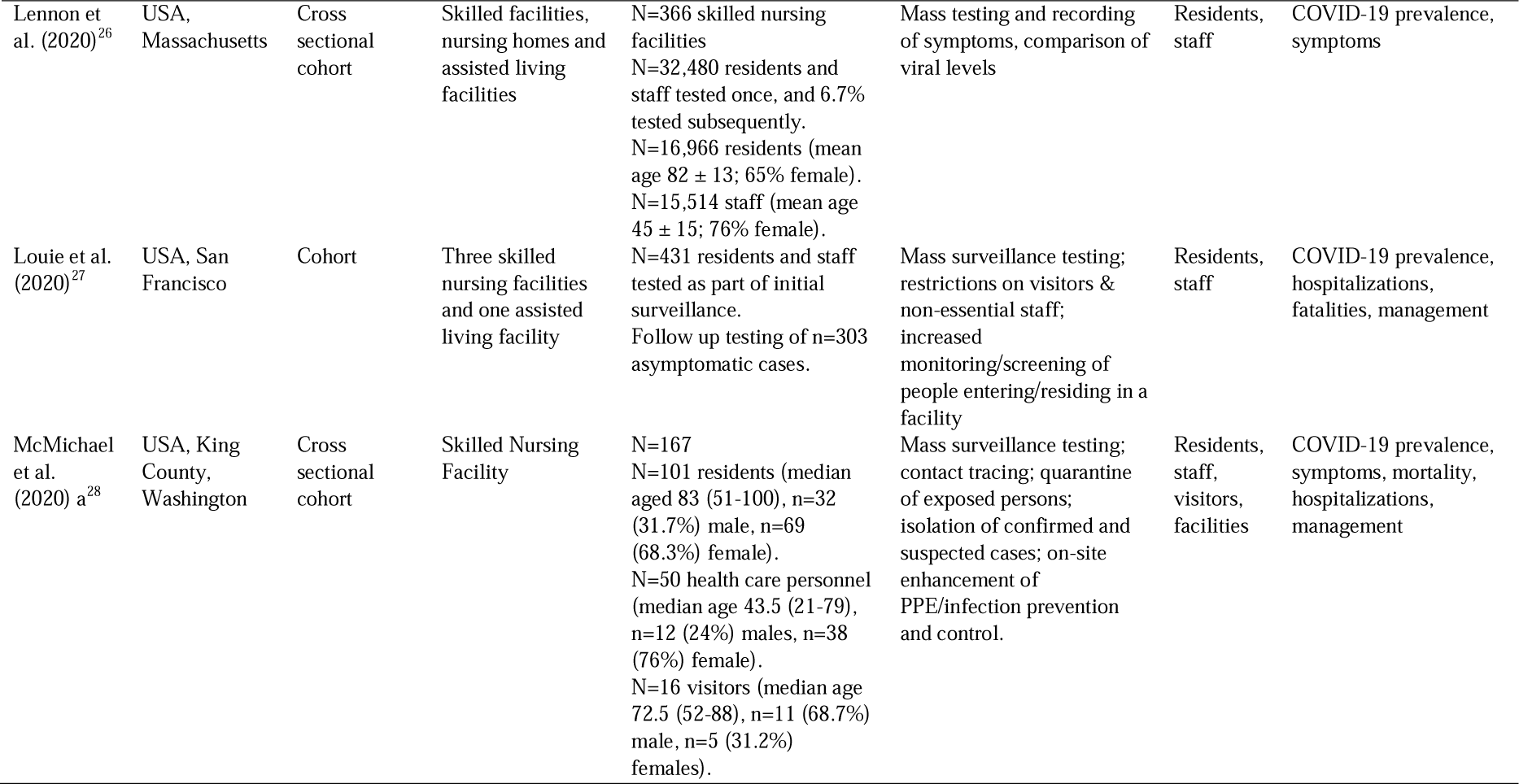
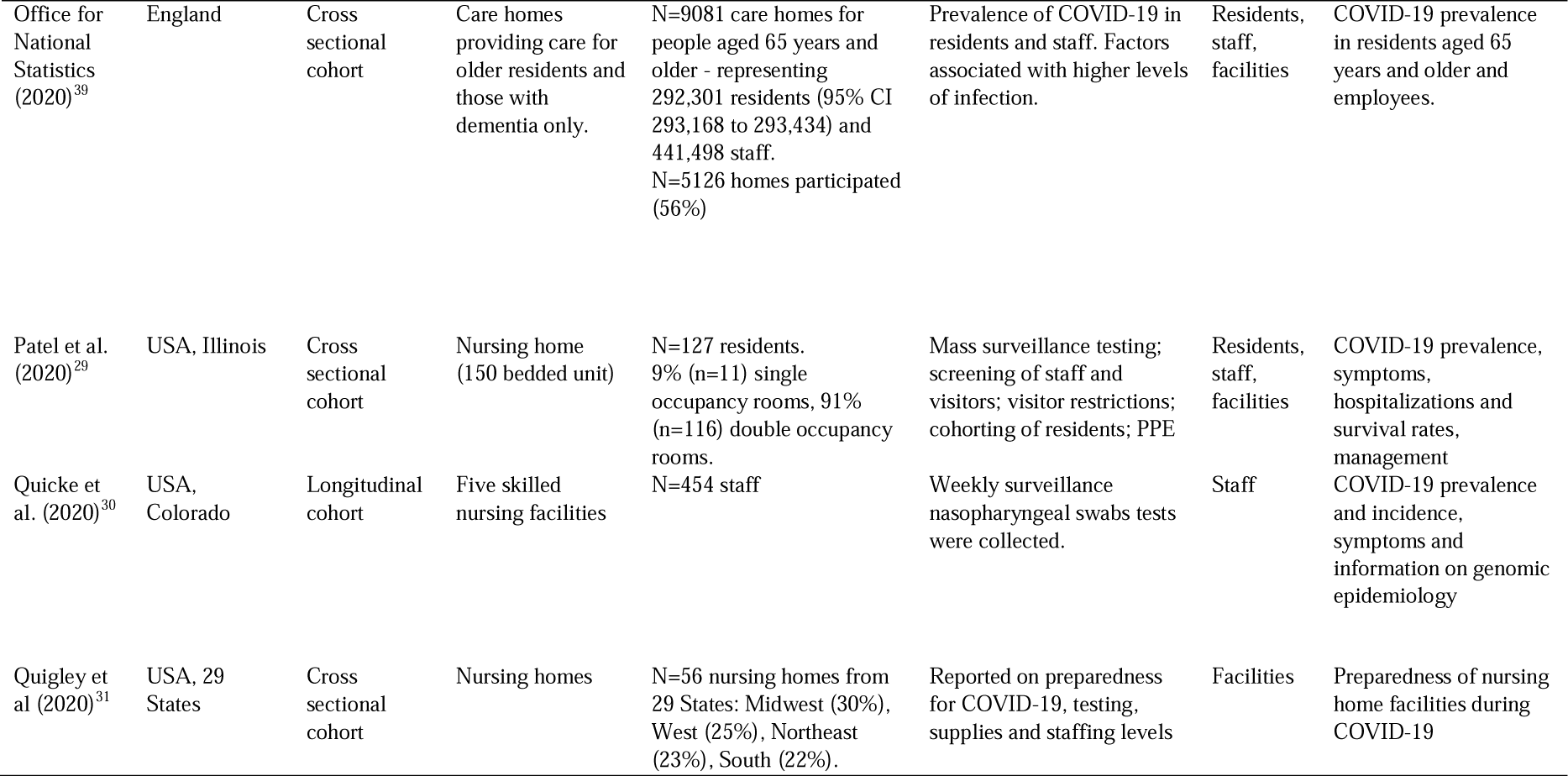
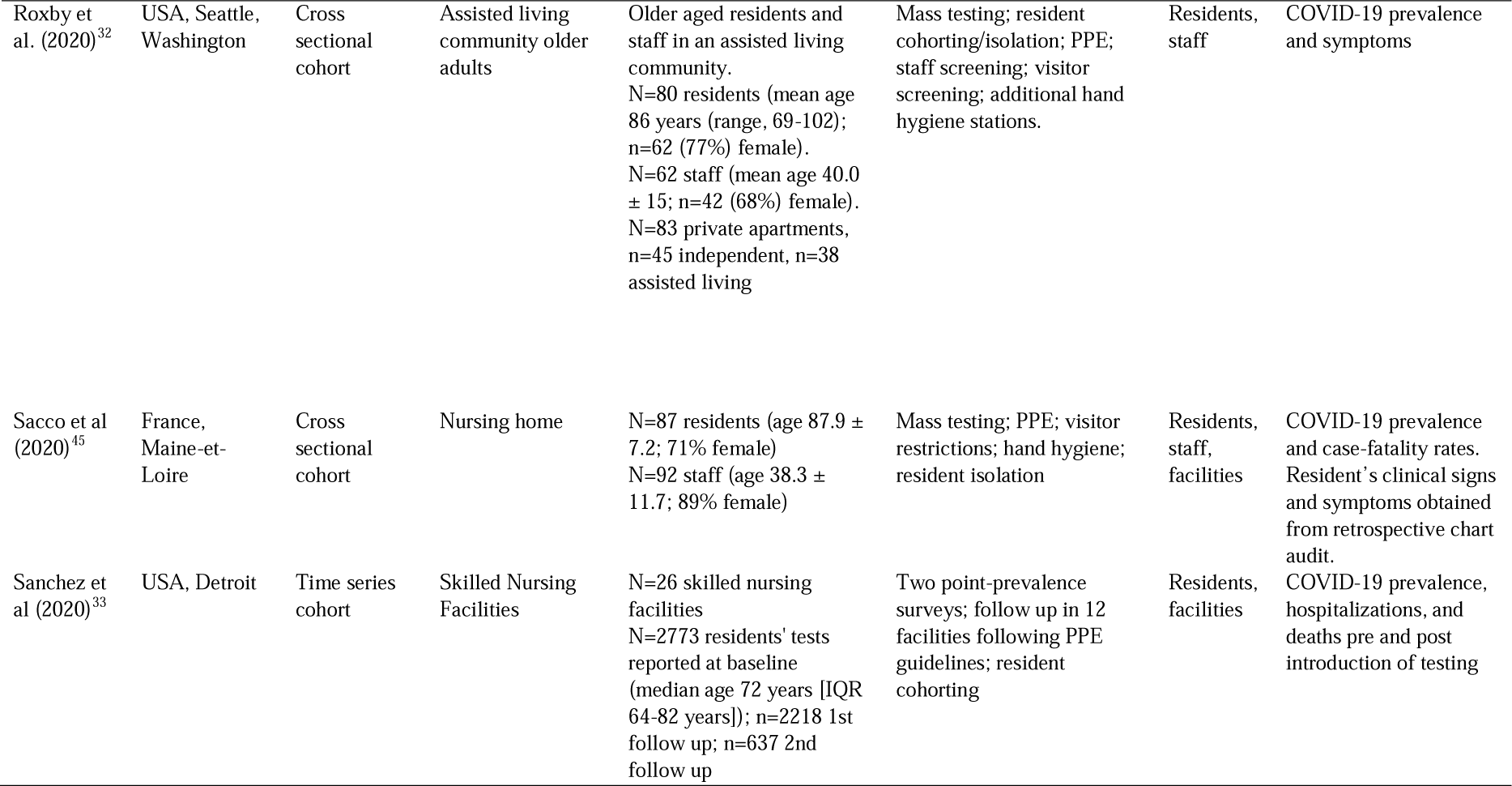
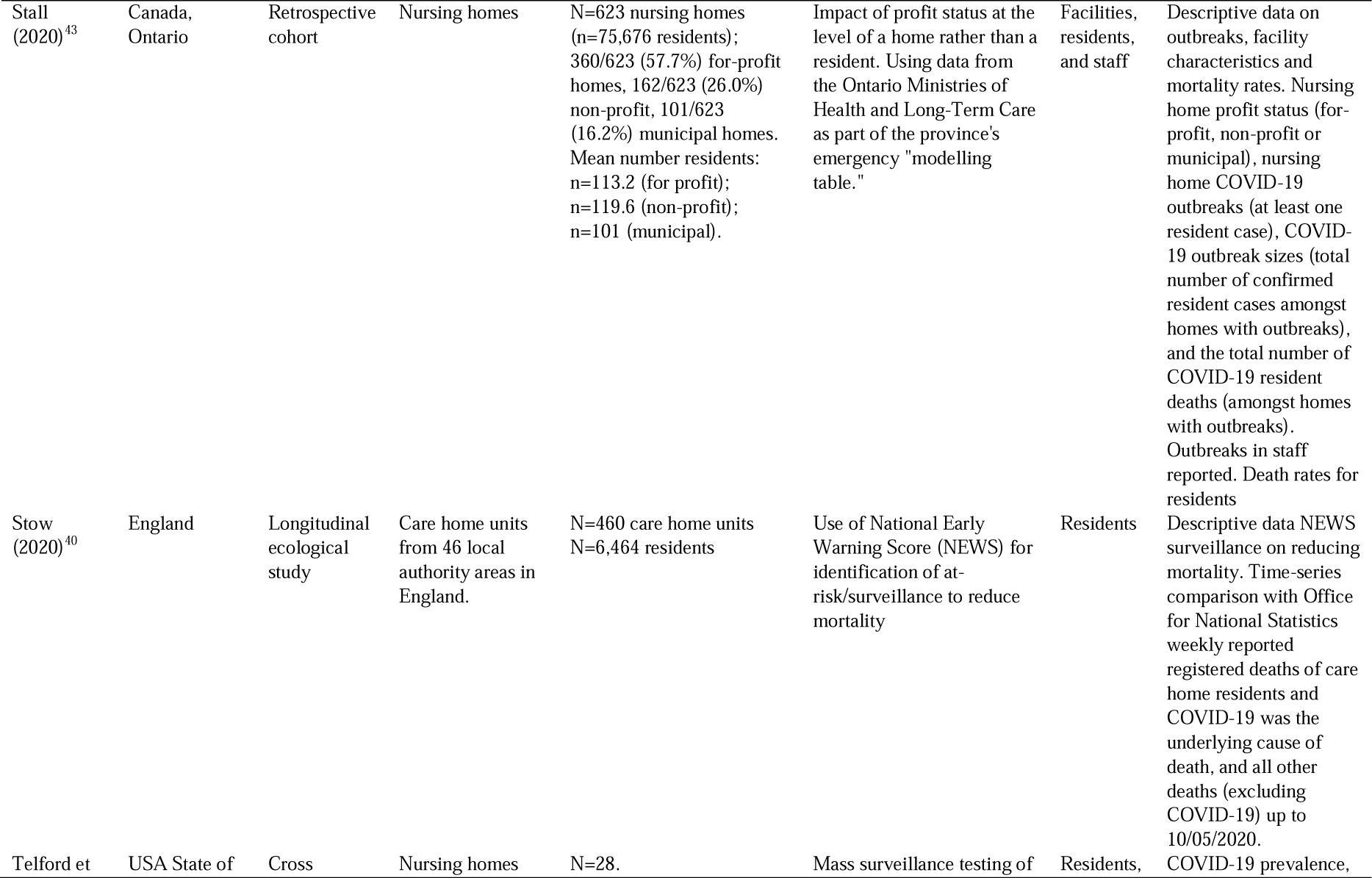
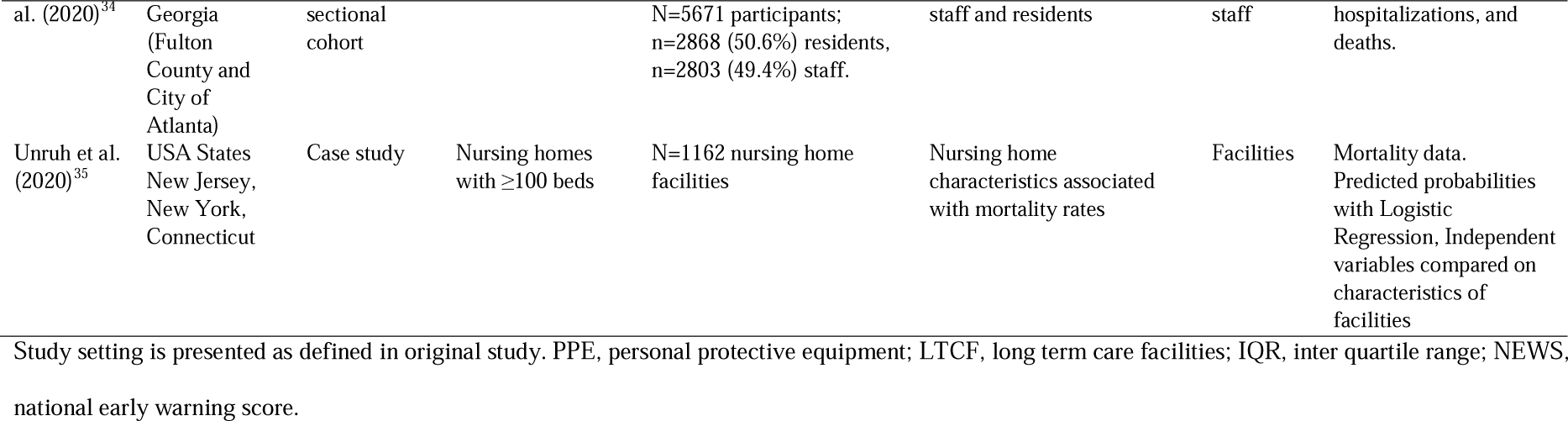
Characteristics of studies including infection control measures

### Infection control measures

Twenty studies report the nature of LTCFs related with outbreaks and transmission of COVID-19 infection (Table 2; ^16,23,28,29,31,33,35-39,41-43,45-47,50-52^). Thirty studies (Table 3a; ^17-29,32-34,37-43,45-50,53^) report evidence of measures to reduce transmission of COVID-19 in long-term residential care facilities for residents, 25 studies (Table 3b; ^17-22,24,26-30,32,34,38,39,42-48,50,53^) report evidence for employee outcomes, and two studies report evidence for visitors (Table 3c; ^28,47^).

**Table 2.**
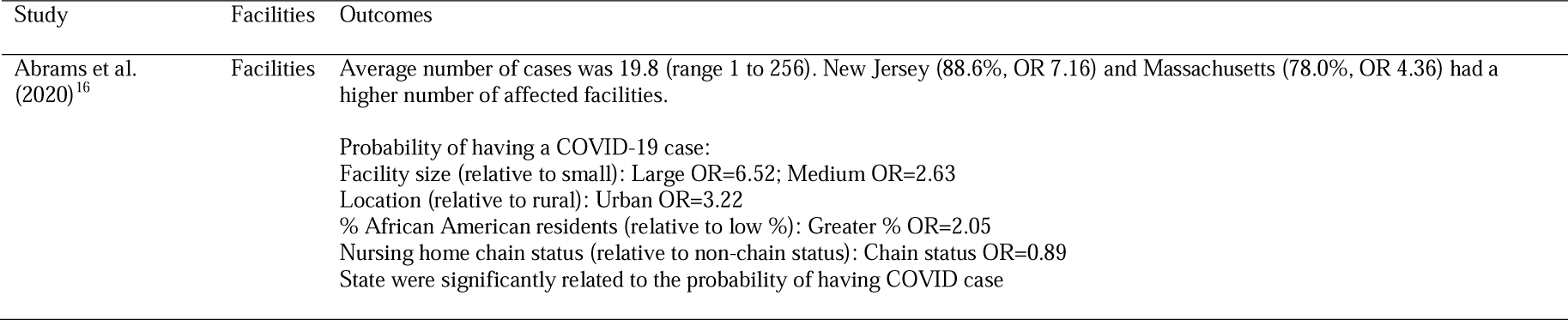
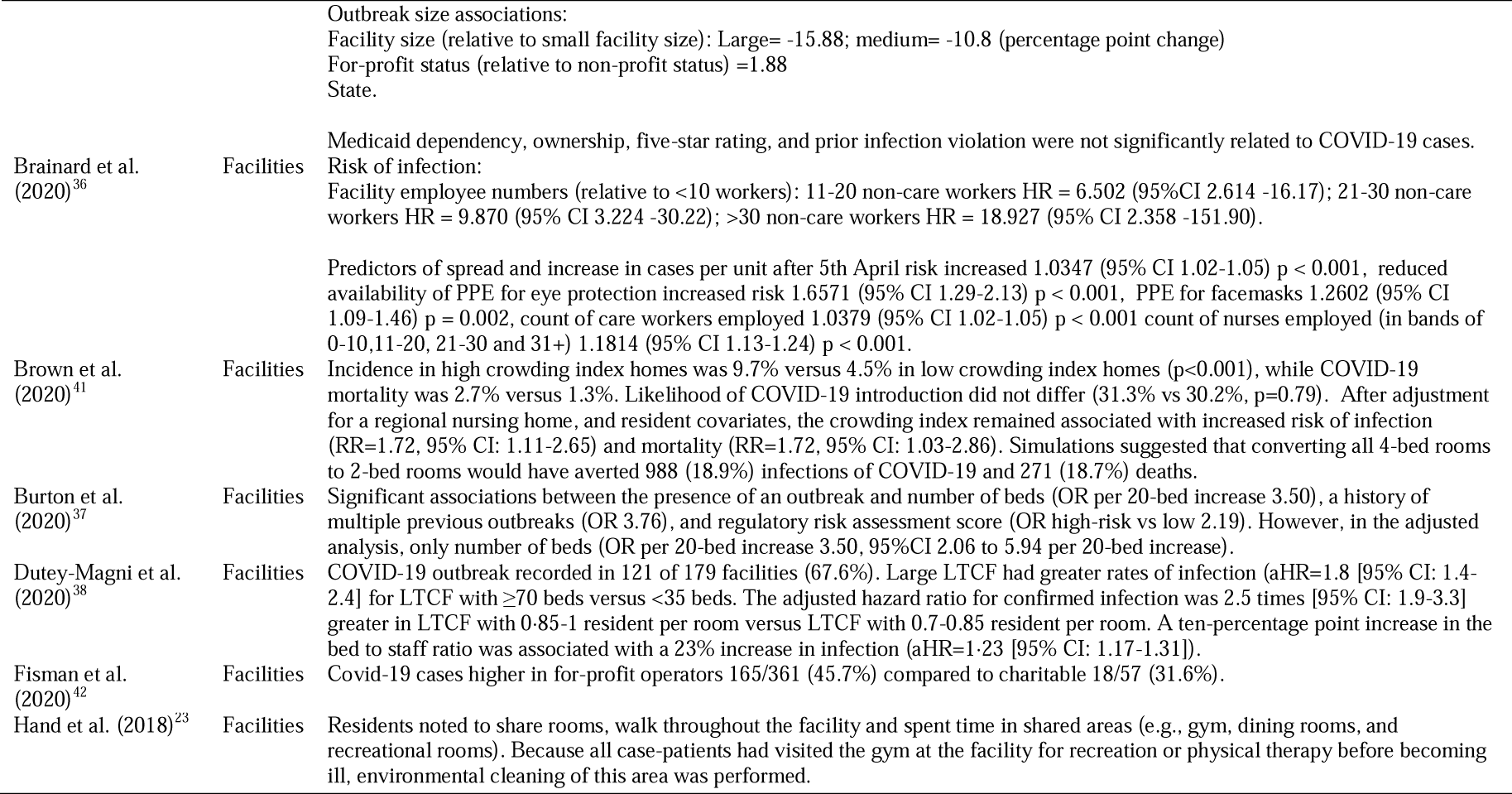
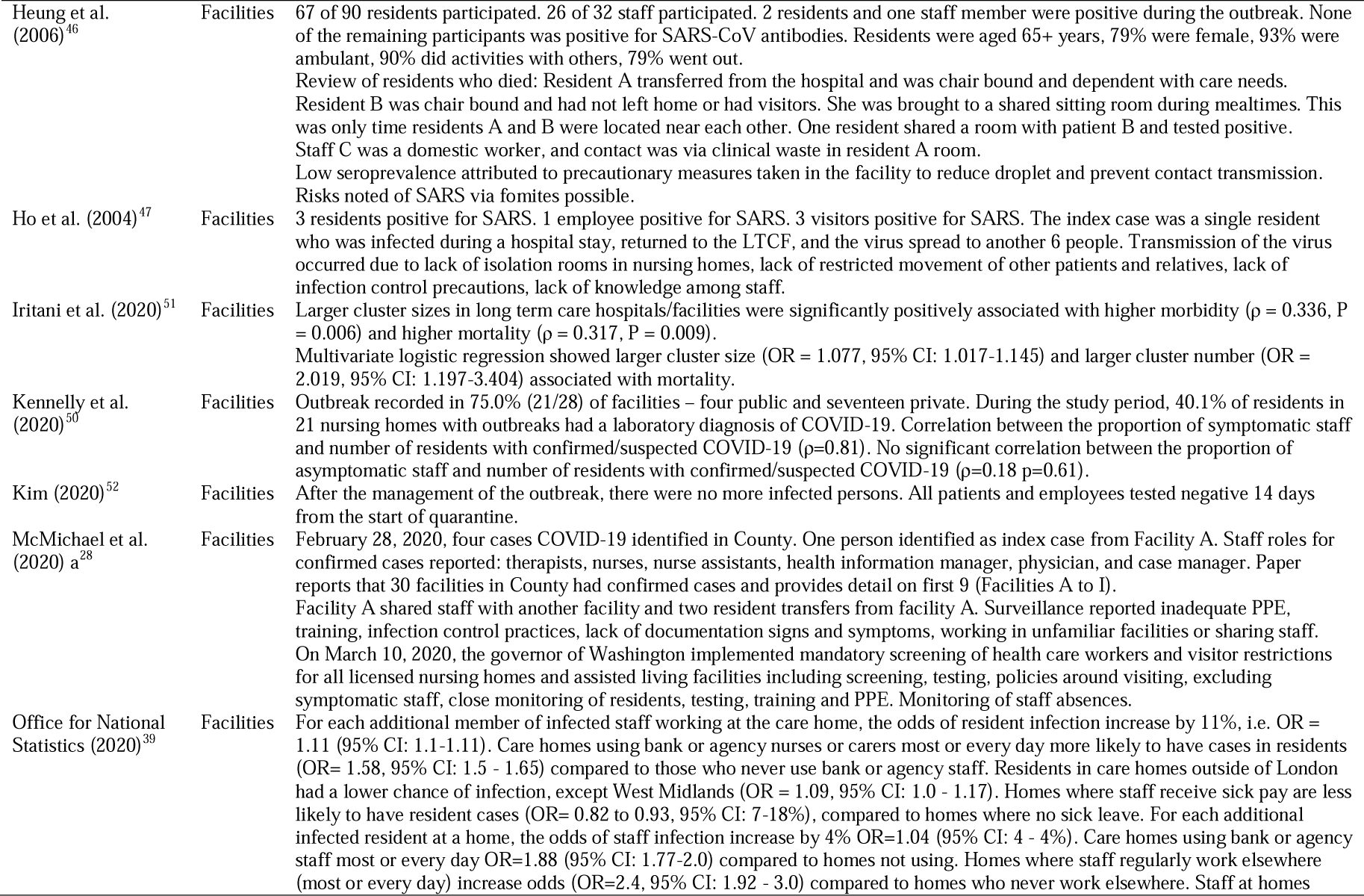
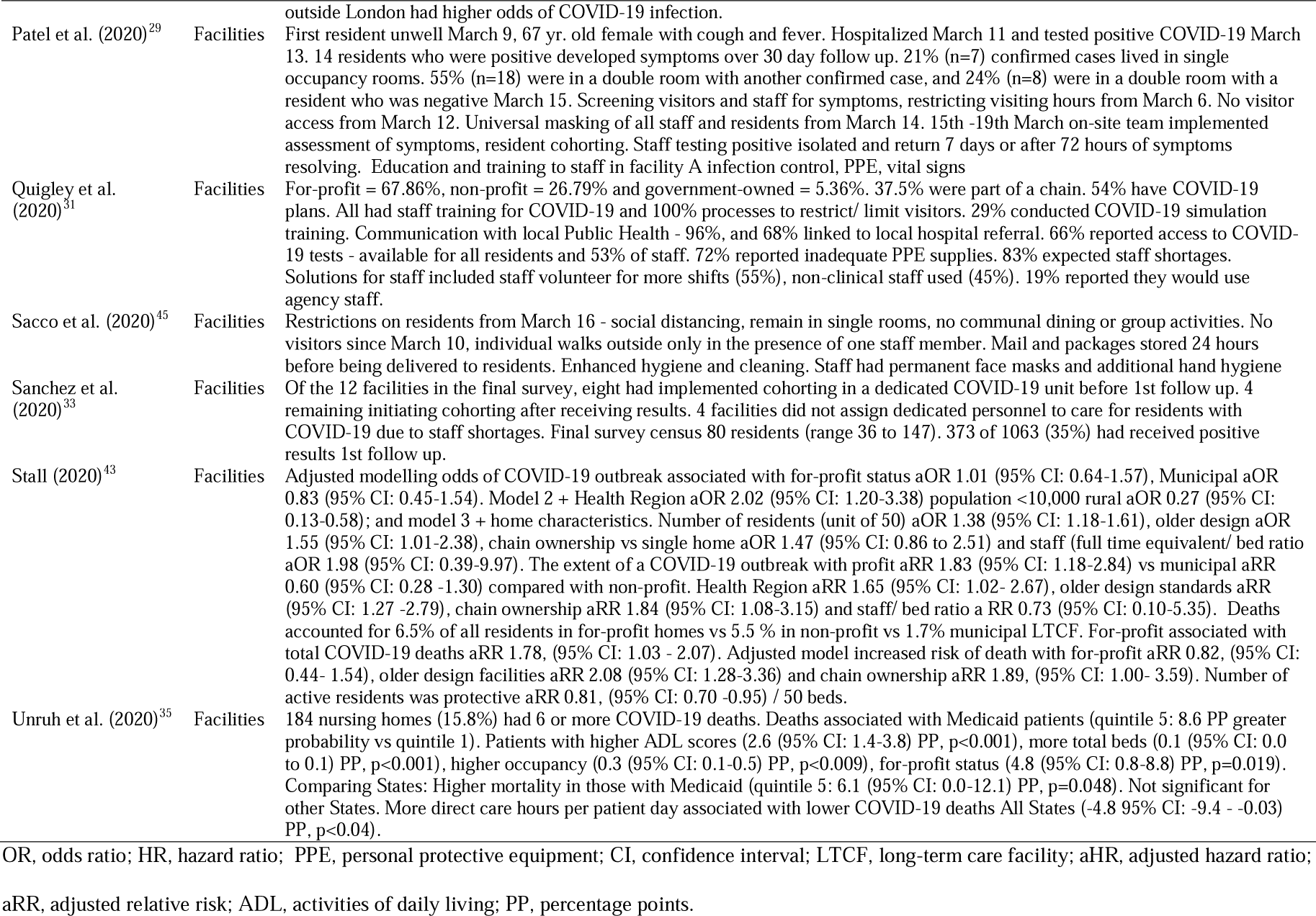
COVID-19 outcomes related to the nature of long term care facilities.

**Table 3a.**
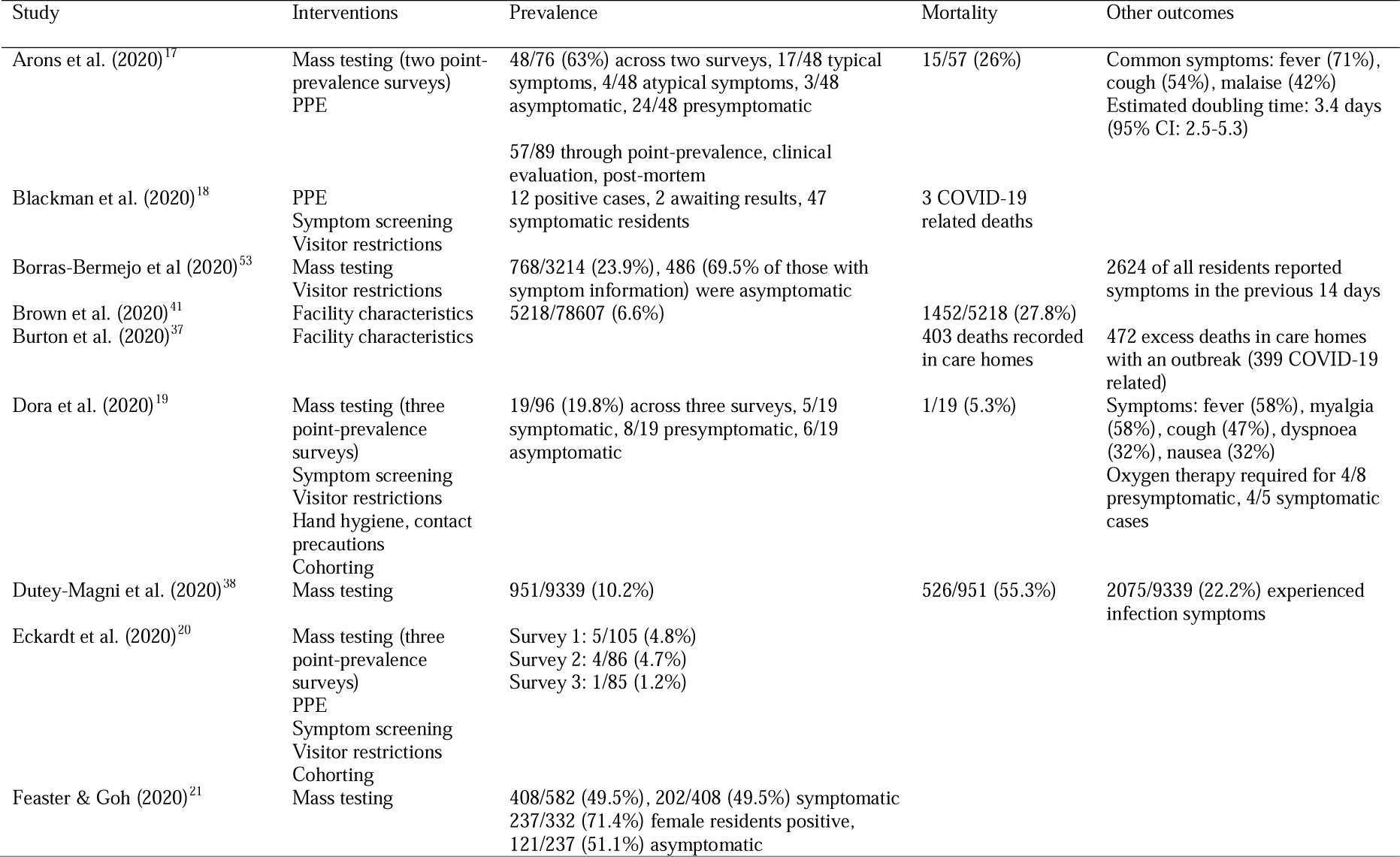
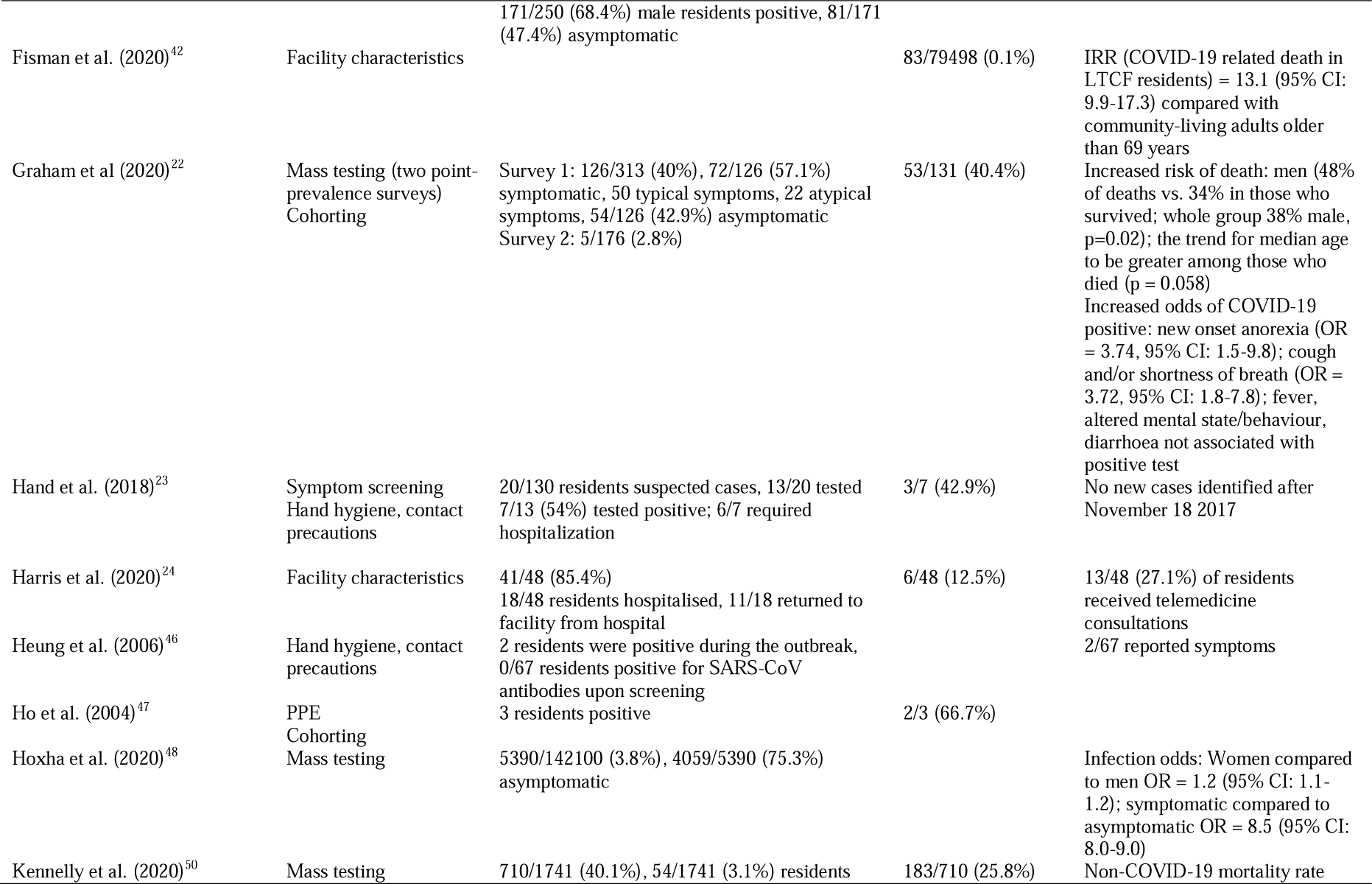
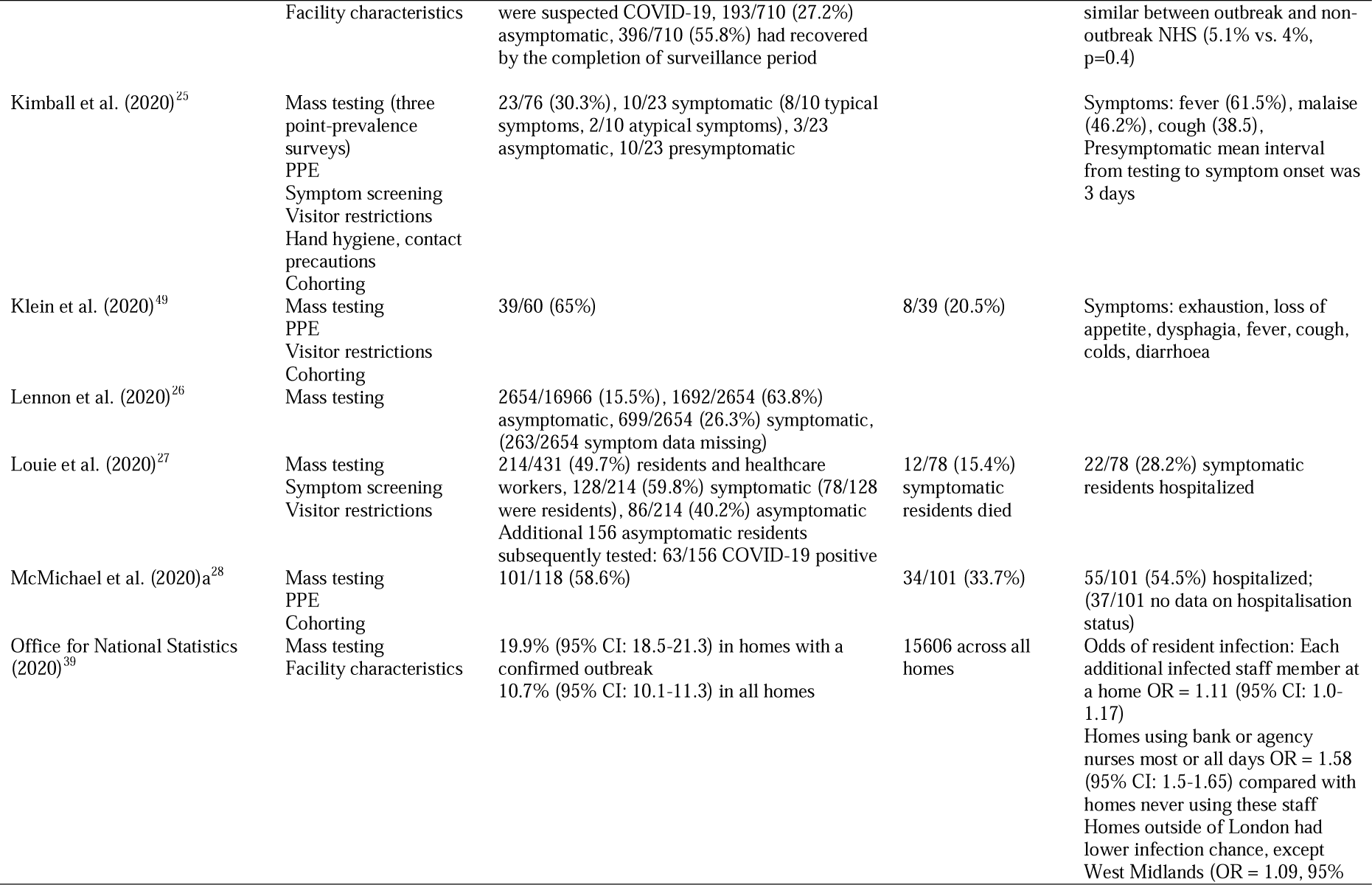
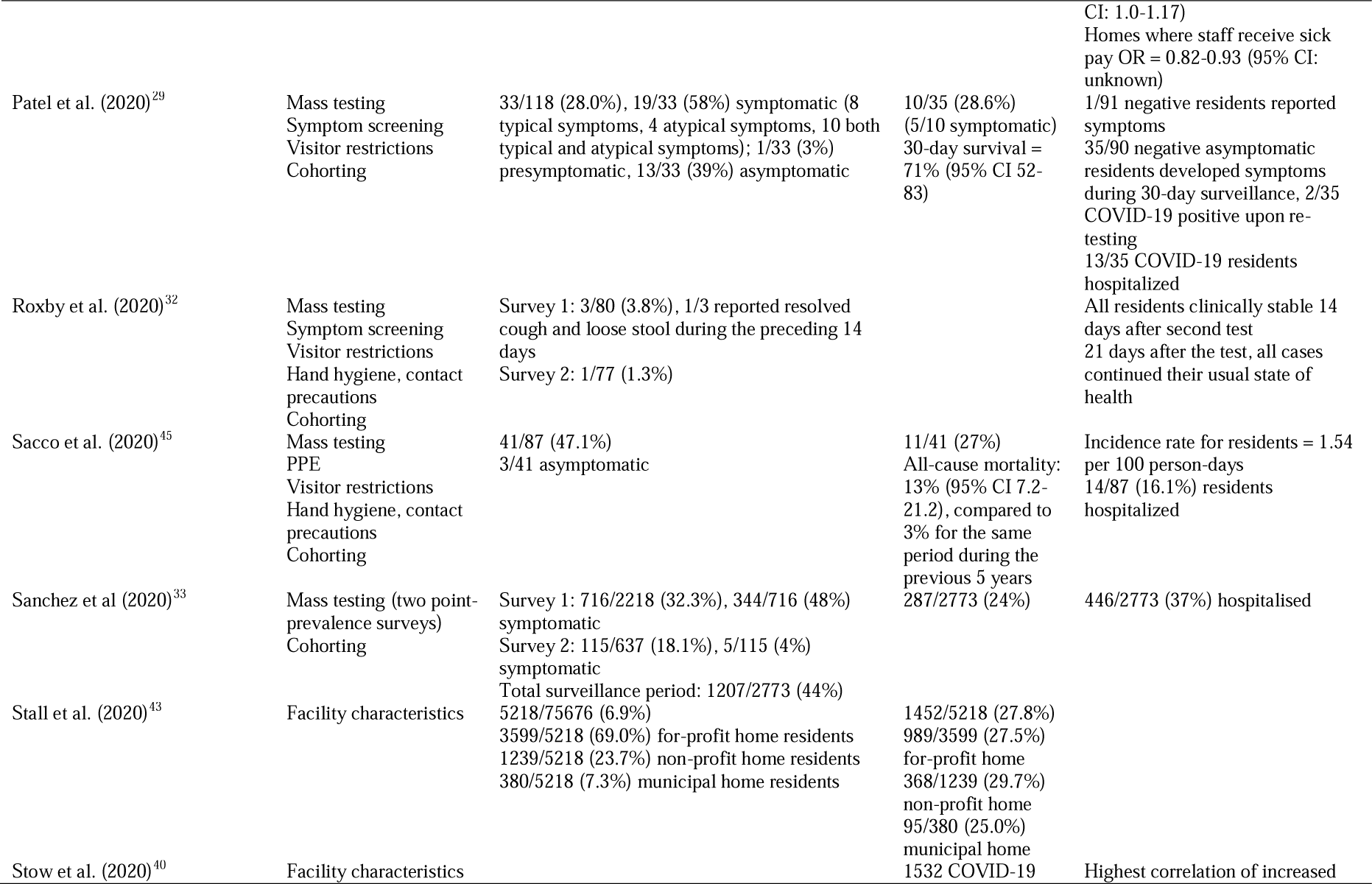
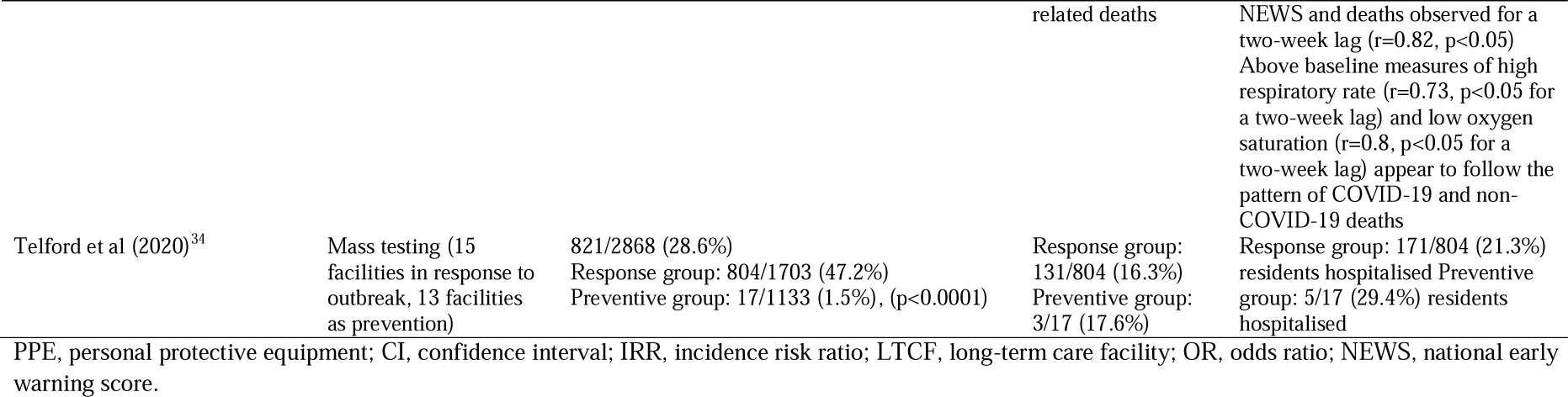
Resident-specific outcomes of strategies implemented in nursing homes

**Table 3b.**
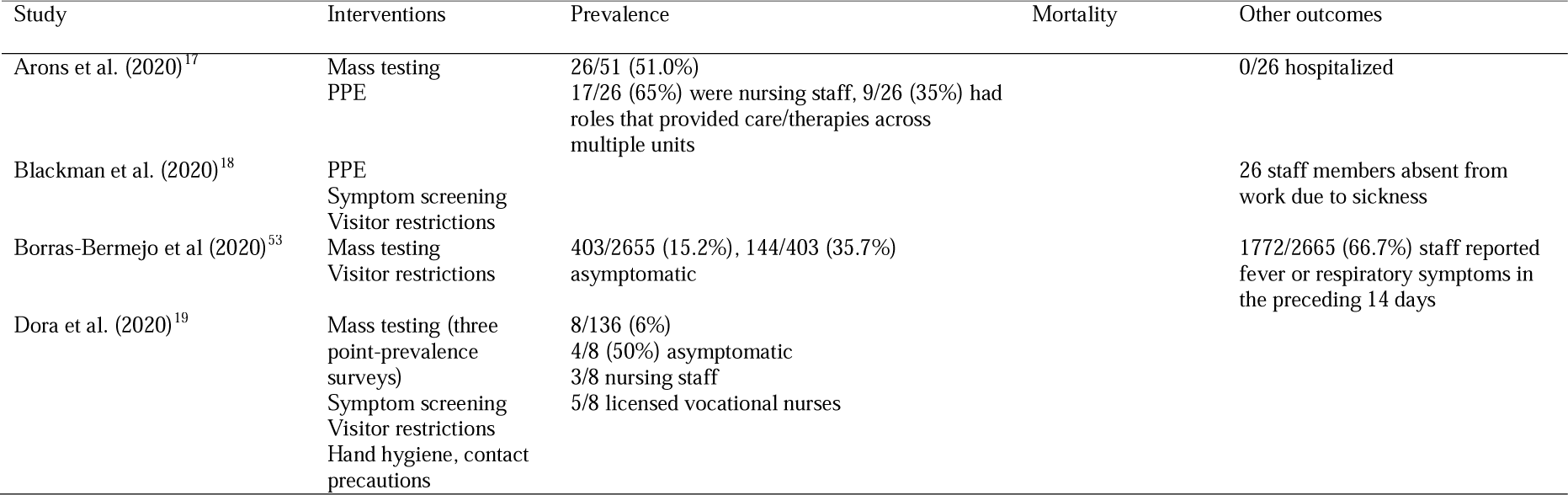
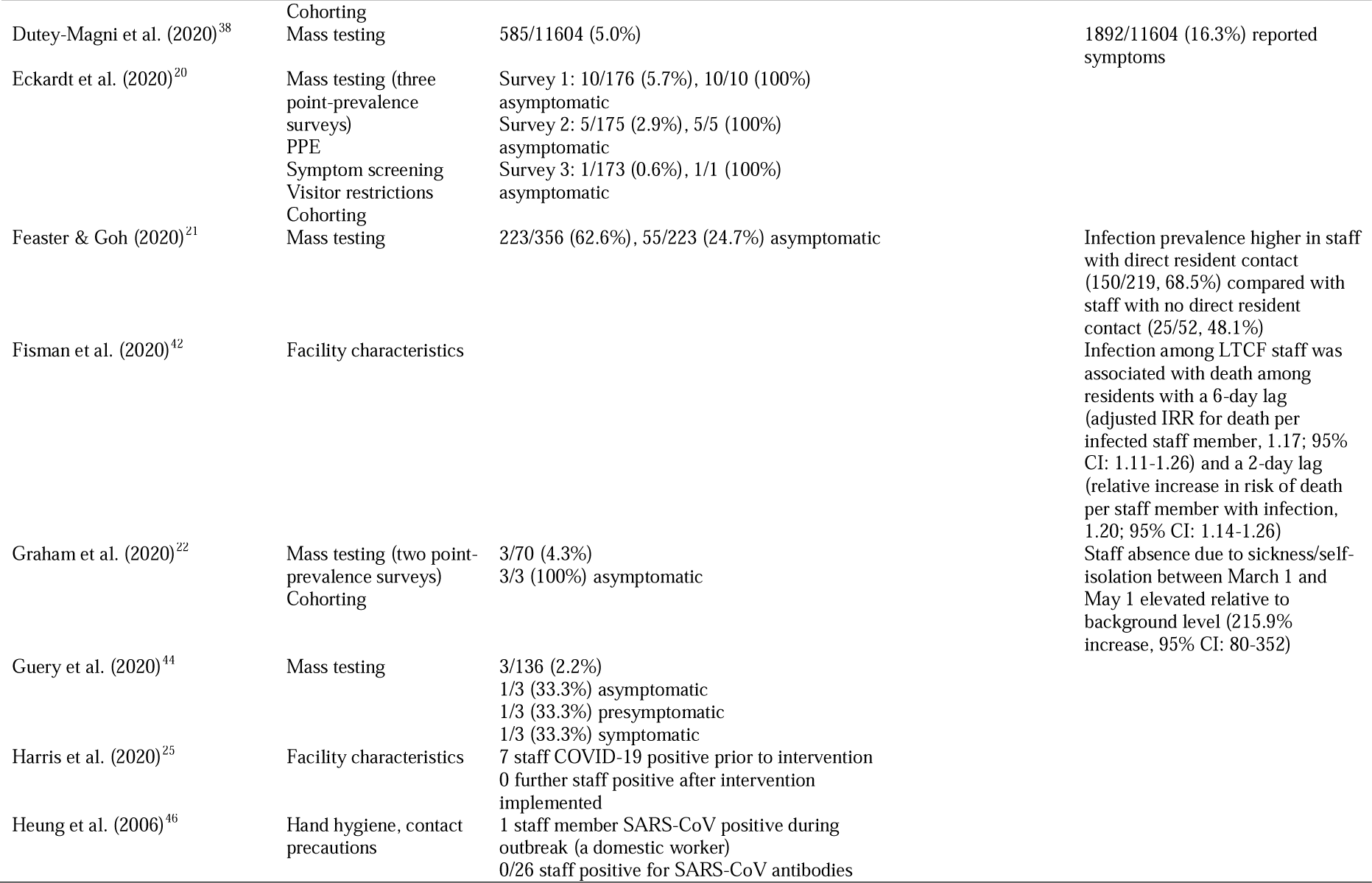
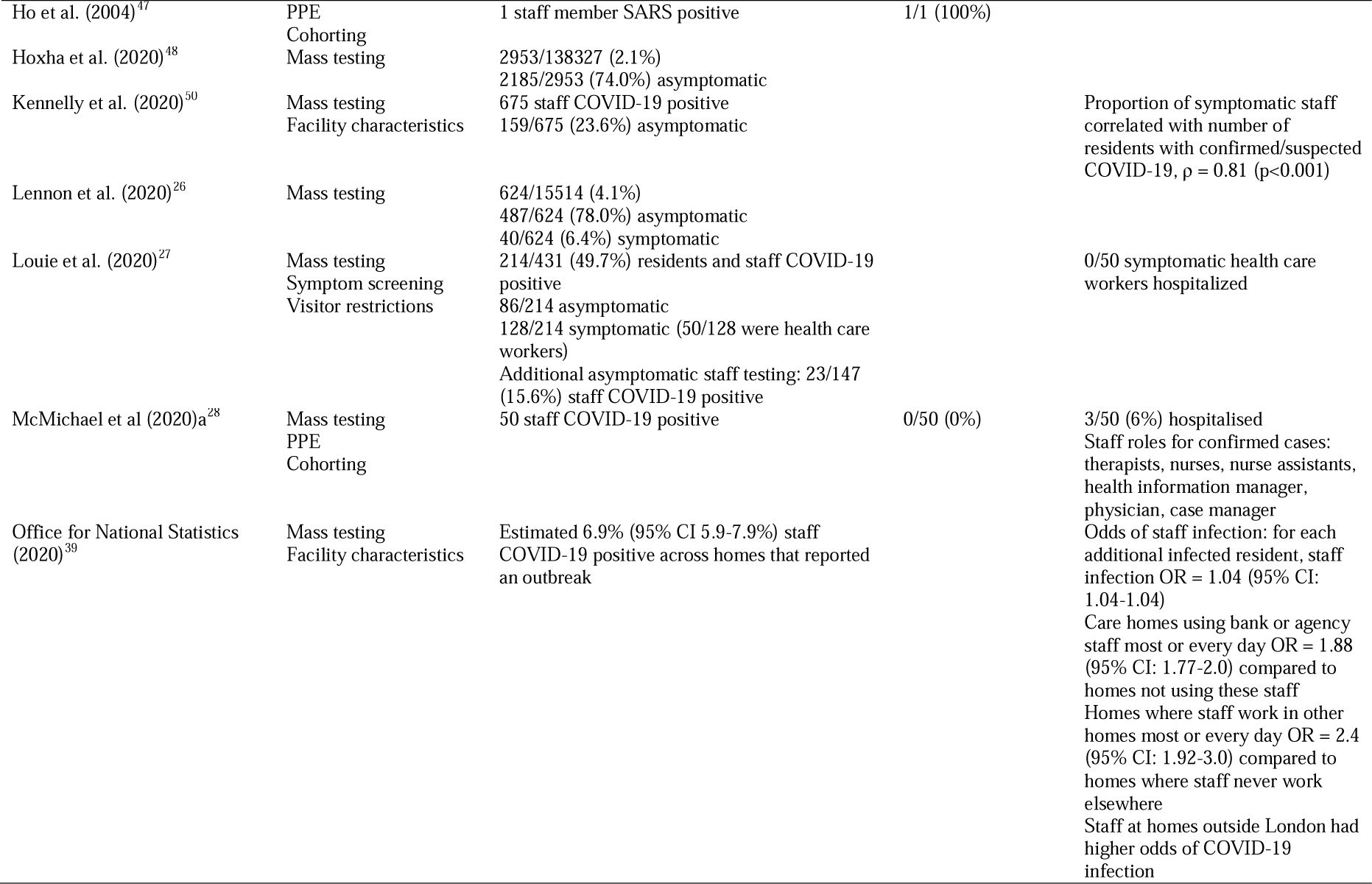
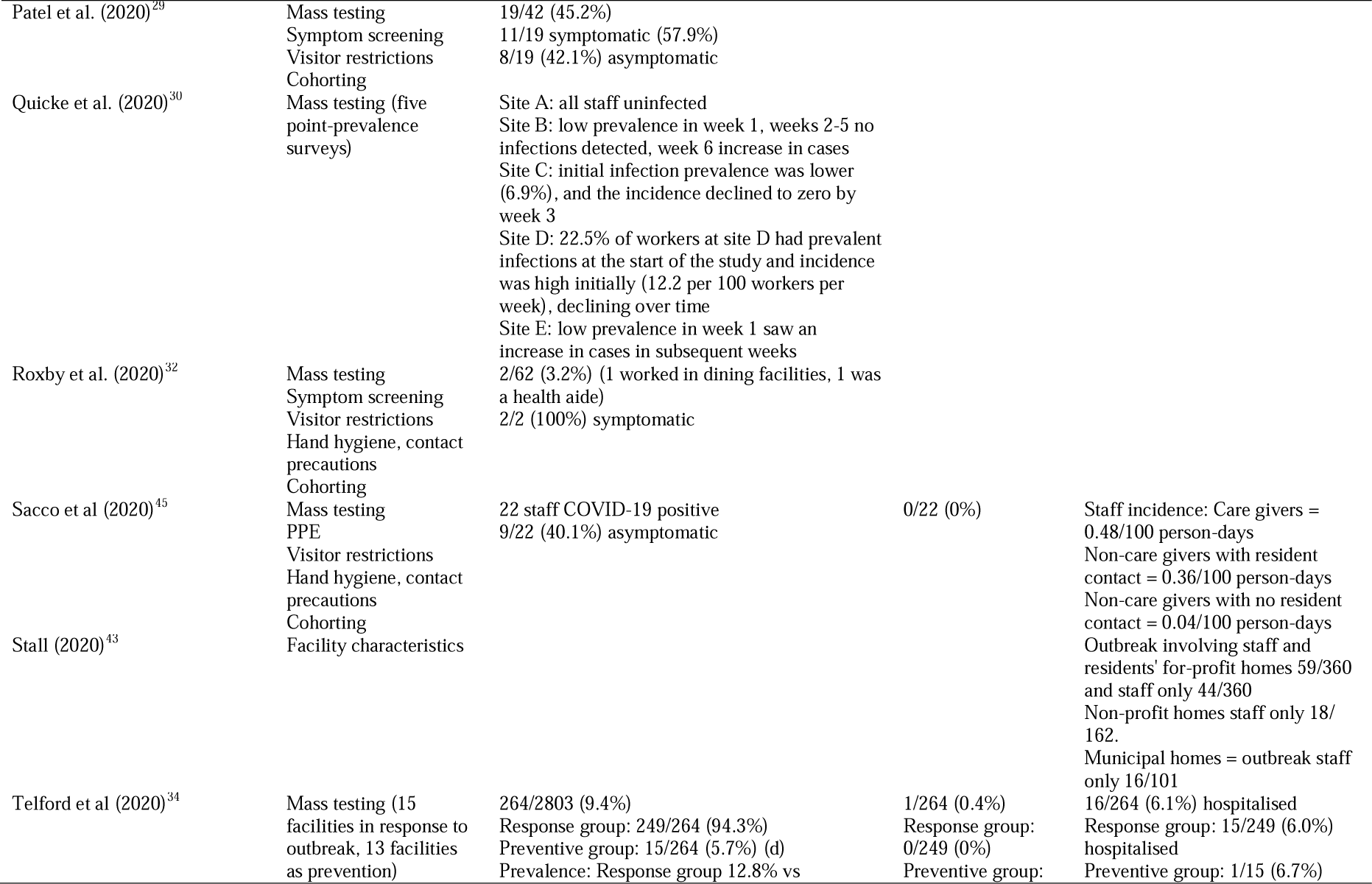
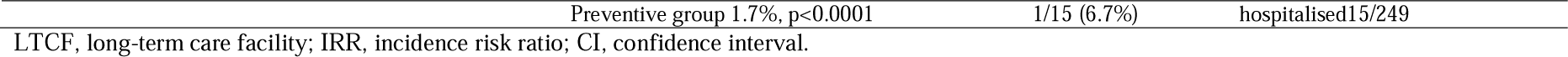
Staff-specific outcomes of strategies to reduce transmission

**Table 3c.**
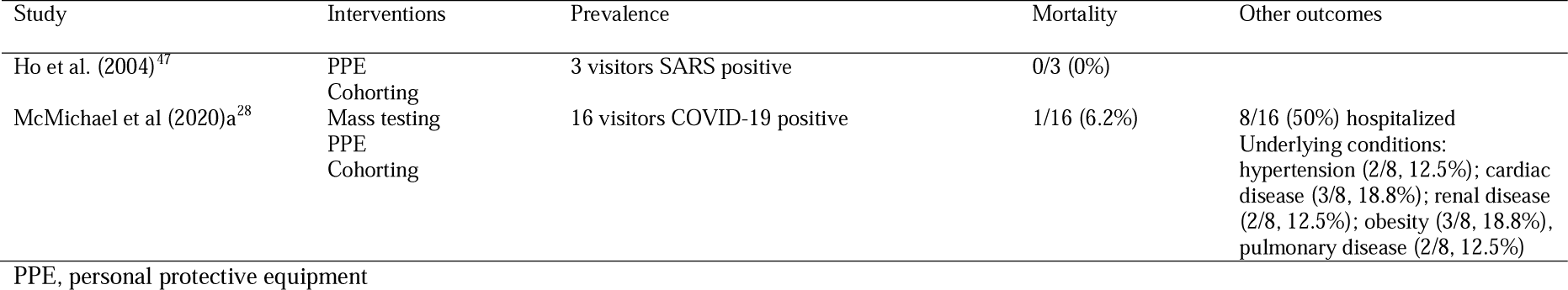
Visitor-specific outcomes following the implementation of strategiesansmission

A variety of infection control measures are described (Tables 1 and 3a-c) including: mass testing/point-prevalence testing (22 studies; ^17,19-22,25-30,32-34,38,39,44,45,48-50,53^), use of personal protective equipment (10 studies; ^17,18,20,25,28,29,32,45,47,49^), screening of residents, staff, or visitors for symptoms (8 studies; ^18-20,23,25,27,29,32^), restrictions on visitor entry (10 studies; ^18-20,25,27,29,32,45,49,53^), hand hygiene and contact and droplet precautions (6 studies; ^19,23,25,32,45,46^), and cohorting/isolation of residents (11 studies; ^19,20,22,25,28,29,32,33,45,47,49^). Thirteen studies examined characteristics of LTCF and their association with COVID-19 infection and risk ^16,24,31,35-37,39-43,51,52^.

### Morbidity and mortality

Morbidity and mortality results from included studies are presented for residents (Table 3a), staff (Table 3b), and visitors (Table 3c). Prevalence of COVID-19 infection was reported in 29 studies, including prevalence in residents (27 studies; ^17-29,32-34,38,39,41,43,45-50,53^) and staff (22 studies; ^17,19-22,24,26-30,32,34,38,39,44-48,50,53^), with 2 studies reporting absolute case numbers in visitors.^28,47^ Prevalence rates ranged from 3.8% in a sample of 2074 LTCF^48^ and 1.2% in the third point-prevalence survey at a single facility^20^ to 85.4% in a single facility that implemented a telemedicine service to limit transmission.^24^ Staff prevalence ranged from 0.6% in a point-prevalence survey in a single facility^20^ to 62.6% in a group of nine LTCF.^21^ One study reported 16 COVID-19 positive visitor cases,^28^ while a study which examined SARS infection following an outbreak in a Hong Kong facility reported 3 positive visitor cases.^47^

The symptom status (symptomatic/presymptomatic/asymptomatic, typical/atypical symptoms) of participants was reported in 16 studies, with resident and staff symptom status reported in 15 ^17-19,21,22,25-27,29,32,33,45,48,50,53^ and 13 studies,^19-22,26,27,29,32,44,45,48,50,53^ respectively. No studies reported symptom status of visitors. The proportion of COVID-19 positive residents presenting with symptoms ranged from 26.3%^19,26^ to 59.8% (a sample of both residents and healthcare workers).^27^ Asymptomatic cases in residents were reported in 13 studies,^17,19,21,22,25-27,29,32,45,48,50,53^ with proportions of COVID-19 positive residents presenting with no symptoms varying from 2.4%^45^ to 75.3%.^48^ Among COVID-19 positive staff, the proportion of symptomatic cases ranged from 6.4%^26^ to 100%,^32^ and asymptomatic cases ranged from 23.6%^50^ to 100%.^20,22^

Mortality results were reported in 22 studies, including information on mortality of residents (22 studies; ^17-19,22-24,27-29,33,34,37-43,45,47,49,50^), staff (4 studies; ^28,34,45,47^), and visitors (2 studies; ^28,47^). Mortality rates in COVID-19 positive residents ranged from 5.3%^19^ to 55.3%.^38^ One study reported a 66.7% death rate in residents who tested positive for the SARS virus.^47^ A study examining the mortality risk in Ontario LTCF reported a death rate of 0.1% across all residents.^42^ Across the three studies which presented mortality results in COVID-19 positive staff, mortality rates were 0%.^28,34,45^ One study presenting mortality rates in a nursing home following a SARS outbreak reported one death of a member of staff.^47^ Mortality rates reported in visitors in two studies was 0%^47^ and 6.2%,^28^ respectively.

### Characteristics of LTCFs on COVID-19 transmission

Numerous facility-specific characteristics were linked with risk of COVID-19 cases (Table 2). These include size of LTCF;^16,37,38,51^ staffing levels and/or use of agency care staff;^28,31,36,38,39,43,50^ part of larger chain of organisations and/or for profit status;^16,31,35,42,43,50^ and related staffing, crowding, or availability of single rooms.^23,29,39,41,43,45-47^

### Quality review

The quality ratings of included studies are presented in Supplementary Table 2. Overall quality of evidence in this review is considered low based on MMAT assessment criteria.

## Discussion

Evidence in this review indicates the impact of COVID-19 on LTCF, demonstrating the vulnerability of this setting. A novel outcome highlights the characteristics of LTCF associated with COVID-19 outbreaks, in addition to reporting the prevalence rates of COVID-19 and associated mortality and morbidity for residents, staff, and visitors. A variety of measures were implemented in LTCF, of which many were instigated locally by facility managers, and others through agile public health policy. Mass testing of residents with or without staff testing was the primary measure used to reduce transmission of COVID-19. This provides objective evidence of infection rates in facilities, and enables application of subsequent measures, including isolation of residents who are infected with re-designation of specific staff to care for them. Repeated point-prevalence testing allows facilities to grasp the spread of the virus along with the impact of their mitigation strategies.

Further measures implemented in facilities echoed public health recommendations to the broader community to limit the spread of the virus. These included guidance on hand hygiene, and contact and droplet precautions. Restricting visitor access to facilities was implemented generally to reduce the likelihood of introducing COVID-19 into LTCF, with assessment of body temperature and symptom screening of staff and visitors on entry.

The prevalence of COVID-19 infection varied throughout included studies, with no distinct pattern emerging between prevention strategies and infection prevalence. Similarly, the mortality rate varied widely among studies and prevention measures. However, patterns emerged regarding associations between facility characteristics and the risk of a COVID-19 outbreak and spread.

The facility size/number of beds was significantly associated with the probability of having a COVID-19 case, and the resulting size of an outbreak. For example, in a sample of 30 US nursing homes, the probability of having a COVID-19 case was increased in medium and large facilities compared with small facilities,^16^ while in 121 UK homes reporting an outbreak, facilities with ≥70 beds had 80% greater infection rates than facilities with <35 beds.^38^ A sample of 623 Canadian nursing homes demonstrated facilities with a high crowding index had more infections and deaths than those with a low crowding index. Simulations conducted suggested nearly 20% of infections and deaths may have been averted by converting all 4-bed rooms into 2-bed rooms.^41^ Similarly, facilities with a greater number of staff, staff who work in multiple facilities, and greater number of infected staff, were also more likely to experience a COVID-19 outbreak.^36,39,50^ However, facilities where staff receive sick leave were shown to be less likely to have positive cases.^39^ Reduced availability of PPE predicted the spread and increase in case number in facilities,^36^ while for-profit status of facilities was commonly identified as increasing the odds of case outbreaks relative to non-profit status.^16,31,35,42,43^

### Quality review

The quality of evidence in this review is technically low, primarily reported from observational studies, expert opinion, reporting of outbreaks and describing the process and management (Supplementary Table 2). Factors associated with lower quality of evidence includes the reliance of self-reporting of symptoms, recall bias, use of datasets which may be incomplete, and use of convenience sampling. However, confirmation of COVID-19 in the majority of studies was via laboratory testing. We did not remove any study following our review of quality and the evidence is consistent with real time reporting of data to learn from outbreaks. The Institute of Medicine (2004)^54^ advocates for early detection of epidemics, effective communication to the public, and promotion of research and development for strategic planning.

### Limitations in the review process

A key strength of this review is that it addresses a knowledge gap and has collated evidence from a broad methodological base to report the measures to reduce transmission of COVID-19 in LTFC and reports characteristics of facilities.

Due to the heterogeneity of studies, meta-analysis was not performed, while the descriptive nature of studies prevents identification of a causative relationship between measures and outcomes. Despite this, the systematic approach to this review has identified the scope of interventions implemented in LTFC to reduce COVID-19 transmission.

Publication bias was minimized with inclusion of pre-published evidence, follow up contacts with authors for early reporting, and through the inclusion of observational study designs. Most studies reported are in English, we translated papers from German and Spanish as part of the assessment and review. Outbreak reports include convenience samples or smaller cohorts of residents in LTCF with limited data reported in brief reports and letters. However, real time reporting of outbreaks provides immediate evidence and shared understanding advocated by the Institute of Medicine.^54^

While the present review builds on a review by Salcher-Konrad, Jhass, Naci, Tan, El-Tawil, Comas-Herrera ^55^, a recent report from WHO,^56^ and from an Irish review report,^57^ data on the role of facilities in the transmission of COVID-19 are reported.

## Conclusion

This novel, rapid review summarises the evidence base to date identifying specific factors for consideration as part of preparedness plans to reduce transmission of COVID-19 outbreaks in LTCF. Future research should incorporate methodologically robust study designs with longer follow up to assess the impact on reducing transmission.

## Supporting information

Supplementary file 1. Search strategy

Supplementary table. Quality ratings results

## Data Availability

Data available upon request

## Author Contributions

CK, KF, and LM designed the study; KF and DS developed the search strategy; DS conducted the literature search; KF and LM screened titles and full texts to select studies, and extracted data; LM, EL, KF, and CK conducted quality ratings; all authors interpreted and synthesised data; all authors were involved in writing. All authors have approved the final version of the manuscript.

