## Supplementary file 1. Search strategy for "A rapid systematic review of measures to protect older people in long term care facilities from COVID-19"

Pubmed

Search #1

“Residential facilit*” OR “Residential aged care” OR Convalescent home* OR “Nursing Home*” OR “Homes for the aged” OR “Housing for the elderly” OR “Skilled nursing facilit*” OR “long term care” OR “Longterm care” OR Home* for the aged OR “Old Age Home*” OR “long-term care” OR "Nursing Homes"[Mesh] OR “long-term care”[MeSH] OR "Residential Facilities"[Mesh] OR "Housing for the Elderly"[Mesh]

213,035 Results

Intervention

Search #2

(“Infection control” OR Infection prevention and control* OR “Patient Safety” OR “Patient harm” OR “Patient risk” OR “Health care Delivery” OR transmission OR body substance isolation* OR physical barrier* OR physical intervention* OR physical protection* OR personal protection* OR person protection* OR BSI OR IPC OR N95 OR ffp1 OR ffp3 OR ffp2 OR transmission* OR contamination* OR shedding OR fomite* OR gap* OR non-pharm intervention* OR non-pharmaceutical intervention* OR Shield OR N99 OR N97 OR Ventilator* OR Space OR spacing or separation OR “Communicable Disease Control” OR "Primary Prevention" OR facemask* OR face mask* OR face-mask* OR "Delivery of Health Care" OR “Disease transmission” OR “Infectious Disease Transmission” OR PPE OR “Personal Protective Equipment” OR mask* OR virucide* OR antivirus agent* OR Handwashing OR “Hand washing” OR “Hand Disinfection” OR “hand hygiene” OR distancing OR distances OR aerosol-generating procedure* OR patient isolation* OR patient isolator* OR person isolator* OR “individual isolation” OR individual isolator* OR filtering face piece* OR face protection* OR face shield* OR face protective device* OR face protective gear* OR eye protection* OR eye shield* OR eye protective device* OR eye protective gear* OR Eye mask* OR airborne precaution* OR droplet precaution* OR safety supply OR safety supplies* OR safety device* OR safety equipment* OR safety measure* OR safety gear* OR protective supply* OR protective supplies* OR protective device* OR protective equipment* OR protective measure* OR protective gear* OR “personal isolation” OR respirator* OR respiratory protection* OR respiratory protective device* OR “respiratory protective supply” OR “respiratory protective supplies” OR “respiratory protective equipment” OR “respiratory protective gear” OR “safely equipped” OR meter OR metre OR foot OR feet OR meters OR metres OR head cover* OR face cover* OR eye cover* OR goggle* OR protective clothing* OR "Infection Control"[Mesh] OR "Personal Protective Equipment"[Mesh] OR "Hand Disinfection"[Mesh] OR "Communicable Disease Control"[Mesh:NoExp] OR "Disease Transmission, Infectious"[Mesh] OR "Primary Prevention"[Mesh] OR "Delivery of Health Care"[Mesh:NoExp] OR "Fomites"[Mesh] OR "Ventilators, Mechanical"[Mesh] OR "Communicable Disease Control"[Mesh] OR "Primary Prevention"[Mesh] OR "Delivery of Health Care"[Mesh] OR "Patient Isolation"[Mesh] OR "Patient Safety"[Mesh] OR "Patient Harm"[Mesh])

5,741,706 results

And

Search #3

(Coronavirus* OR “Corona virus” OR Betacoronavirus or Beta-coronavirus OR Corona* OR coronaviral OR coronavirdae OR coronavirida OR coronaviridae OR coronaviridea OR coronaviridiae OR coronavirinae OR coronavirion OR coronavirions OR coronaviroses OR coronavirous OR coronavirues OR coronaviruscpe OR coronaviruse OR coronaviruses OR coronaviruslike OR coronaviser OR coronaviurs OR coronaviuses OR coronavrius OR coronavvirus OR COVIDOR SARS OR SARS-CoV OR “Middle East respiratory syndrome” OR MERS OR MERS-CoV OR “Severe Acute Respiratory Syndrome” OR “severe acute respiratory pneumonia outbreak” OR 2019-nCoV OR nCoV OR COVID-2019 OR “COVID 2019” OR cov2 OR Covid19 OR COVID-19 OR COVID 19 OR SARS-CoV* OR coronaviridae OR "corona virus" OR "SARS-CoV-2" OR "sars cov2" OR "SARS-CoV-19" OR 2019nCoV OR "SARS-CoV" OR SARSCOV2 OR "2019 coronavirus" OR "SARS2" OR "2019 corona virus" OR covid19 OR "novel corona virus" OR "new corona virus" OR "novel coronavirus" OR "new coronavirus" OR “coronavirus infection” OR "nouveau coronavirus" OR "COVID-19" [Supplementary Concept] OR "severe acute respiratory syndrome coronavirus 2" [Supplementary Concept] OR "Coronavirus Infections"[Mesh] OR "Coronavirus"[Mesh] OR "Middle East Respiratory Syndrome Coronavirus"[Mesh] OR "Coronavirus Infections"[Mesh] OR "SARS Virus"[Mesh] OR "Betacoronavirus"[Mesh])

595,661 results

Search #4 = #2 AND #3 116,217 results

Outcomes

Search #5

Mortality OR “Death rate*” OR “Mortality Rate*” OR Morbidity OR “Risk of Infection” OR “infection risk” OR "Mortality"[Mesh:NoExp] OR "Morbidity"[Mesh]

3,204,107 results

Search #6 = #1 AND #4 AND #5

EMBASE

Search #1

“Residential facilit*” OR “Residential aged care” OR “Convalescent home*” OR “Nursing Home*” OR “Homes for the aged” OR “Housing for the elderly” OR “Skilled nursing facilit*” OR “long term care” OR “Longterm care” OR “Home* for the aged” OR “Old Age Home*” OR “long-term care” OR 'residential home'/exp OR 'nursing home'/exp OR 'home for the aged'/exp OR 'skilled nursing facility'/exp OR 'long term care'/de

212,416 results

Intervention

Search #2

(“Infection control” OR “Infection prevention and control*” OR “Patient Safety” OR “Patient harm” OR “Patient risk” OR “body substance isolation*” OR “physical barrier*” OR “physical intervention*” OR “physical protection*” OR “personal protection*” OR “person protection*” OR BSI OR IPC OR N95 OR ffp1 OR ffp3 OR ffp2 OR transmission* OR contamination* OR shedding OR fomite* OR gap* OR “non-pharm intervention*” OR “non-pharmaceutical intervention*” OR Shield OR N99 OR N97 OR Ventilator* OR Space OR spacing OR separation OR “Communicable Disease Control” OR "Primary Prevention" OR facemask* OR face-mask* OR “face mask*” OR "Delivery of Health Care" OR “Health Care Delivery” OR “Disease transmission” OR “Infectious Disease Transmission” OR PPE OR “Personal Protective Equipment” OR mask* OR virucide* OR antivirus agent* OR Handwashing OR “Hand washing” OR “Hand Disinfection” OR “hand hygiene” OR distancing OR distances OR “aerosol-generating procedure*” OR “patient isolation*” OR “patient isolator*” OR “person isolator*” OR “individual isolation” OR “individual isolator*” OR “filtering face piece*” OR “face protection*” OR “face shield*” OR “face protective device*” OR “face protective gear*” OR “eye protection*” OR “eye shield*” OR “eye protective device*” OR “eye protective gear*” OR “Eye mask*” OR “airborne precaution*” OR “droplet precaution*” OR “safety supply” OR “safety supplies*” OR “safety device*” OR “safety equipment*” OR “safety measure*” OR “safety gear*” OR “protective supply*” OR “protective supplies*” OR “protective device*” OR “protective equipment*” OR “protective measure*” OR “protective gear*” OR “personal isolation” OR respirator* OR “respiratory protection*” OR “respiratory protective device*” OR “respiratory protective supply” OR “respiratory protective supplies” OR “respiratory protective equipment” OR “respiratory protective gear” OR “safely equipped” OR meter OR metre OR foot OR feet OR meters OR metres OR “head cover*” OR “face cover*” OR “eye cover*” OR goggle* OR “protective clothing*” OR 'infection control'/exp OR 'patient safety'/exp OR 'disease transmission'/exp OR 'contamination'/exp OR 'shedding'/exp OR 'fomite'/exp OR 'shield'/exp OR 'ventilator'/exp OR 'space'/exp OR 'separation'/exp OR 'communicable disease control'/exp OR 'primary prevention'/exp OR 'face mask'/exp OR 'health care delivery'/exp OR 'protective equipment'/exp OR 'mask'/exp OR 'antivirus agent'/exp OR 'hand washing'/exp OR 'patient isolation'/exp OR 'face shield'/exp OR 'eye protective device'/exp OR 'ventilator'/exp OR 'respiratory protection'/exp OR 'goggle'/exp OR 'protective clothing'/exp)

6,030,646 results

And

Search #3

(Coronavirus* OR “Corona virus” OR Betacoronavirus or Beta-coronavirus OR Corona* OR coronaviral OR coronavirdae OR coronavirida OR coronaviridae OR coronaviridea OR coronaviridiae OR coronavirinae OR coronavirion OR coronavirions OR coronaviroses OR coronavirous OR coronavirues OR coronaviruscpe OR coronaviruse OR coronaviruses OR coronaviruslike OR coronaviser OR coronaviurs OR coronaviuses OR coronavrius OR coronavvirus OR COVIDOR SARS OR SARS-CoV OR “Middle East respiratory syndrome” OR MERS OR MERS-CoV OR “Severe Acute Respiratory Syndrome” OR “severe acute respiratory pneumonia outbreak” OR 2019-nCoV OR nCoV OR COVID-2019 OR “COVID 2019” OR cov2 OR Covid19 OR COVID-19 OR COVID 19 OR SARS-CoV* OR "SARS-CoV-2" OR "sars cov2" OR "SARS-CoV-19" OR 2019nCoV OR "SARS-CoV" OR SARSCOV2 OR "2019 coronavirus" OR "SARS2" OR "2019 corona virus" OR covid19 OR "novel corona virus" OR "new corona virus" OR "novel coronavirus" OR "new coronavirus" OR “coronavirus infection” OR "nouveau coronavirus" OR 'Coronavirinae'/exp OR 'Betacoronavirus'/exp OR 'severe acute respiratory syndrome'/exp OR 'covid 19'/exp OR 'Coronavirus infection'/exp)

45,801 results

Search #4 = #2 AND #3 27,921 results

Outcomes

Search #5

Mortality OR “Death rate*” OR “Mortality Rate*” OR Morbidity OR “Risk of Infection” OR “Infection risk” OR 'mortality'/exp OR 'mortality rate'/exp OR 'morbidity'/exp OR 'infection risk'/exp

1,862,861results

Search #6 = #1 AND #4 AND #5

CINAHL

Search #1

“Residential facilit*” OR “Residential aged care” OR “Convalescent home*” OR “Nursing Home*” OR “Homes for the aged” OR “Housing for the elderly” OR “Skilled nursing facilit*” OR “long term care” OR “Longterm care” OR “Home* for the aged” OR “Old Age Home*” OR “long-term care” OR (MH "Residential Facilities") OR (MH "Nursing Homes+") OR (MH "Housing for the Elderly") OR (MH "Long Term Care")

83,231 results

Intervention

Search #2

(“Infection control” OR “Infection prevention and control*” OR “Patient Safety” OR “Patient harm” OR “Patient risk” OR “body substance isolation*” OR “physical barrier*” OR “physical intervention*” OR “physical protection*” OR “personal protection*” OR “person protection*” OR BSI OR IPC OR N95 OR ffp1 OR ffp3 OR ffp2 OR transmission* OR contamination* OR shedding OR fomite* OR gap* OR “non-pharm intervention*” OR “non-pharmaceutical intervention*” OR Shield OR N99 OR N97 OR Ventilator* OR Space OR spacing OR separation OR “Communicable Disease Control” OR "Primary Prevention" OR facemask* OR face-mask* OR “face mask*” OR "Delivery of Health Care" OR “Health Care Delivery” OR “Disease transmission” OR “Infectious Disease Transmission” OR PPE OR “Personal Protective Equipment” OR mask* OR virucide* OR antivirus agent* OR Handwashing OR “Hand washing” OR “Hand Disinfection” OR “hand hygiene” OR distancing OR distances OR “aerosol-generating procedure*” OR “patient isolation*” OR “patient isolator*” OR “person isolator*” OR “individual isolation” OR “individual isolator*” OR “filtering face piece*” OR “face protection*” OR “face shield*” OR “face protective device*” OR “face protective gear*” OR “eye protection*” OR “eye shield*” OR “eye protective device*” OR “eye protective gear*” OR “Eye mask*” OR “airborne precaution*” OR “droplet precaution*” OR “safety supply” OR “safety supplies*” OR “safety device*” OR “safety equipment*” OR “safety measure*” OR “safety gear*” OR “protective supply*” OR “protective supplies*” OR “protective device*” OR “protective equipment*” OR “protective measure*” OR “protective gear*” OR “personal isolation” OR respirator* OR “respiratory protection*” OR “respiratory protective device*” OR “respiratory protective supply” OR “respiratory protective supplies” OR “respiratory protective equipment” OR “respiratory protective gear” OR “safely equipped” OR meter OR metre OR foot OR feet OR meters OR metres OR “head cover*” OR “face cover*” OR “eye cover*” OR goggle* OR “protective clothing*” OR (MH "Infection Control") OR (MH "Handwashing") OR (MH "Patient Safety") OR (MH "Disease Transmission+") OR (MH "Microbial Contamination") OR (MH "Ventilators, Mechanical") OR (MH "Masks") OR (MH "Health Care Delivery+") OR (MH "Protective Devices+") OR (MH "Patient Isolation+")

917,391 results

And

Search #3

(Coronavirus* OR “Corona virus” OR Betacoronavirus or Beta-coronavirus OR Corona* OR coronaviral OR coronavirdae OR coronavirida OR coronaviridae OR coronaviridea OR coronaviridiae OR coronavirinae OR coronavirion OR coronavirions OR coronaviroses OR coronavirous OR coronavirues OR coronaviruscpe OR coronaviruse OR coronaviruses OR coronaviruslike OR coronaviser OR coronaviurs OR coronaviuses OR coronavrius OR coronavvirus OR COVIDOR SARS OR SARS-CoV OR “Middle East respiratory syndrome” OR MERS OR MERS-CoV OR “Severe Acute Respiratory Syndrome” OR “severe acute respiratory pneumonia outbreak” OR 2019-nCoV OR nCoV OR COVID-2019 OR “COVID 2019” OR cov2 OR Covid19 OR COVID-19 OR COVID 19 OR SARS-CoV* OR OR "SARS-CoV-2" OR "sars cov2" OR "SARS-CoV-19" OR 2019nCoV OR "SARS-CoV" OR SARSCOV2 OR "2019 coronavirus" OR "SARS2" OR "2019 corona virus" OR covid19 OR "novel corona virus" OR "new corona virus" OR "novel coronavirus" OR "new coronavirus" OR “coronavirus infection” OR "nouveau coronavirus" OR (MH "Coronavirus+") OR (MH "Coronaviridae Infections+") )

141,416 results

Search #4 = #2 AND #3 15,251 results

Outcomes

Search #5

Mortality OR “Death rate*” OR “Mortality Rate*” OR Morbidity OR “Risk of Infection” OR “Infection risk” OR (MH "Mortality+") OR (MH "Morbidity+")

501,502 results

Search #6 = #1 AND #4 AND #5

Cochrane library

#1	MeSH descriptor: [Coronavirus Infections] explode all trees	179

#2	MeSH descriptor: [Coronavirus] explode all trees	18

#3	Coronavirus OR “Corona virus” OR Betacoronavirus or Beta-coronavirus OR Corona OR coronaviral OR coronavirdae OR coronavirida OR coronaviridae OR coronaviridea OR coronaviridiae OR coronavirinae OR coronavirion OR coronavirions OR coronaviroses OR coronavirous OR coronavirues OR coronaviruscpe OR coronaviruse OR coronaviruses OR coronaviruslike OR coronaviser OR coronaviurs OR coronaviuses OR coronavrius OR coronavvirus OR COVIDOR SARS OR SARS-CoV OR “Middle East respiratory syndrome” OR MERS OR MERS-CoV OR “Severe Acute Respiratory Syndrome” OR “severe acute respiratory pneumonia outbreak” OR nCoV OR COVID-2019 OR “COVID 2019” OR cov2 OR Covid19 OR COVID-19 OR COVID 19 OR SARS-CoV* OR coronaviridae OR "corona virus" OR "SARS-CoV-2" OR "sars cov2" OR "SARS-CoV-19" OR 2019nCoV OR "SARS-CoV" OR SARSCOV2 OR "2019 coronavirus" OR "SARS2" OR "2019 corona virus" OR covid19 OR "novel corona virus" OR "new corona virus" OR "novel coronavirus" OR "new coronavirus" OR “coronavirus infection” OR "nouveau coronavirus"	1173

#4	#1 OR #2 OR #3	1173

#5	“Infection control” OR Infection prevention and control* OR “Patient Safety” OR “Patient harm” OR “Patient risk” OR “Health care Delivery” OR transmission OR body substance isolation* OR physical barrier* OR physical intervention* OR physical protection* OR personal protection* OR person protection* OR BSI OR IPC OR N95 OR ffp1 OR ffp3 OR ffp2 OR transmission* OR contamination* OR shedding OR fomite* OR gap* OR non-pharm intervention* OR non-pharmaceutical intervention* OR Shield OR N99 OR N97 OR Ventilator* OR Space OR spacing or separation OR “Communicable Disease Control” OR "Primary Prevention" OR facemask* OR face mask* OR face-mask* OR "Delivery of Health Care" OR “Disease transmission” OR “Infectious Disease Transmission” OR PPE OR “Personal Protective Equipment” OR mask* OR virucide* OR antivirus agent* OR Handwashing OR “Hand washing” OR “Hand Disinfection” OR “hand hygiene” OR distancing OR distances OR aerosol-generating procedure* OR patient isolation* OR patient isolator* OR person isolator* OR “individual isolation” OR individual isolator* OR filtering face piece* OR face protection* OR face shield* OR face protective device* OR face protective gear* OR eye protection* OR eye shield* OR eye protective device* OR eye protective gear* OR Eye mask* OR airborne precaution* OR droplet precaution* OR safety supply OR safety supplies* OR safety device* OR safety equipment* OR safety measure* OR safety gear* OR protective supply* OR protective supplies* OR protective device* OR protective equipment* OR protective measure* OR protective gear* OR “personal isolation” OR respirator* OR respiratory protection* OR respiratory protective device* OR “respiratory protective supply” OR “respiratory protective supplies” OR “respiratory protective equipment” OR “respiratory protective gear” OR “safely equipped” OR meter OR metre OR foot OR feet OR meters OR metres OR head cover* OR face cover* OR eye cover* OR goggle* OR protective clothing*	300480

#6	MeSH descriptor: [Infection Control] explode all trees	1147

#7	MeSH descriptor: [Personal Protective Equipment] explode all trees	2284

#8	MeSH descriptor: [Hand Disinfection] explode all trees	378

#9	MeSH descriptor: [Communicable Disease Control] explode all trees	4791

#10	MeSH descriptor: [Disease Transmission, Infectious] explode all trees	856

#11	MeSH descriptor: [Primary Prevention] explode all trees	4005

#12	MeSH descriptor: [Delivery of Health Care] explode all trees	44666

#13	MeSH descriptor: [Fomites] explode all trees	9

#14	MeSH descriptor: [Ventilators, Mechanical] explode all trees	264

#15	MeSH descriptor: [Patient Isolation] explode all trees	51

#16	MeSH descriptor: [Patient Safety] explode all trees	580

#17	MeSH descriptor: [Patient Harm] explode all trees	3

#18	#5 OR #6 OR #7 OR #8 OR #9 OR #10 OR #11 OR #12 OR #13 OR #14 OR #15 OR #16 OR #17	336372

#19	#18 AND #4	651

#20	residential facilit* OR residential aged care OR Convalescent home OR Nursing home* OR Homes for the aged OR Housing for the elderly OR Skilled nursing facilit* OR Long term care OR Longterm care OR Home* for the aged OR old age home OR Long-term care	121379

#21	MeSH descriptor: [Long-Term Care] explode all trees	1112

#22	MeSH descriptor: [Nursing Homes] explode all trees	1314

#23	MeSH descriptor: [Residential Facilities] explode all trees	1711

#24	MeSH descriptor: [Housing for the Elderly] explode all trees	39

#25	#20 OR #21 OR #22 OR #23 OR #24	121450

#26	Mortality OR “Death rate*” OR “Mortality Rate*” OR Morbidity OR “Risk of Infection” OR “infection risk”	111129

#27	MeSH descriptor: [Mortality] explode all trees	12838

#28	MeSH descriptor: [Morbidity] explode all trees	14392

#29	#26 OR #27 OR #28	124060

#30	#19 AND #25 AND #29

Medrxiv

"((COVID-19 OR SARS-CoV-2) And ("Infection control")) AND (Mortality) AND ("nursing homes")"
