## Supplementary table. Quality ratings results for "A rapid systematic review of measures to protect older people in long term care facilities from COVID-19"

| Supplemental Table 2. Quality Review | | | | | | | | | | | | | | |
| --- | --- | --- | --- | --- | --- | --- | --- | --- | --- | --- | --- | --- | --- | --- |
|  |  | ***S1*** | ***S2*** | ***3.1*** | ***3.2*** | ***3.3*** | ***3.4*** | ***3.5*** | ***4.1*** | ***4.2*** | ***4.3*** | ***4.4*** | ***4.5*** | ***Comments*** |
| Abrams (2020) | Quantitative descriptive | Y | Y |  |  |  |  |  | Y | CT | Y | CT | Y |  |
| Arons (2020) | Quantitative descriptive | Y | Y |  |  |  |  |  | Y | Y | Y | N | Y |  |
| Blackman 2020 | Quantitative descriptive | Y | Y |  |  |  |  |  | Y | N | Y | CT | Y* | *Data very limited to descriptive statistics (counts) |
| Borras-Bermejo 2020 | Quantitative descriptive | Y | Y |  |  |  |  |  | Y | CT | Y | N | Y* | *Data minimal descriptive statistics. Reported as a brief letter. |
| Brainard+ (2020) | Non-randomised | Y | Y | CT | Y | Y | CT | Y |  |  |  |  |  |  |
| Brown (2020) + | Non-randomised | Y | Y | Y | Y | Y | N | Y | Y | Y | Y | Y | Y |  |
| Burton (2020) + | Quantitative descriptive | Y | Y |  |  |  |  |  | Y | Y | Y | Y | Y |  |
| Dora (2020) | Quantitative descriptive | Y | Y |  |  |  |  |  | Y | Y | Y | Y | Y* | *Data reporting descriptive data from an outbreak (counts and percentages) |
| Dutey-Magni (2020) + | Non-randomised | Y | Y | Y | Y | Y | N | Y |  |  |  |  |  |  |
| Eckhardt (2020) | Quantitative descriptive | Y | Y |  |  |  |  |  | Y | Y | Y | Y | Y* | *Limited descriptive data (point prevalence data, counts &percentages) |
| Feaster (2020) | Quantitative descriptive | Y | Y |  |  |  |  |  | Y | Y | Y | Y | Y |  |
| Fisman (2020b) | Non-randomised | Y | Y | Y | Y | Y | N | Y |  |  |  |  |  |  |
| Graham (2020) | Quantitative descriptive | Y | Y |  |  |  |  |  | Y | N | Y | Y | Y |  |
| Guery (2020) | Quantitative descriptive | Y | Y |  |  |  |  |  | Y | Y | Y | Y | Y* | *Limited descriptive data reported. Outbreak reported as a published letter. |
| Hand (2018) | Quantitative descriptive | Y | CT |  |  |  |  |  |  |  |  |  |  | Research letter reporting minimal data. |
| Harris (2020) | Quantitative descriptive | Y | Y |  |  |  |  |  | Y | Y | Y | Y | Y* | *Data limited to descriptive statistics |
| Heung (2006) | Quantitative descriptive | Y | Y |  |  |  |  |  | Y | Y | Y | N | Y* | *Limited descriptive data |
| Ho (2003) | Quantitative descriptive | Y | CT |  |  |  |  |  |  |  |  |  |  | Report of conference symposium. Limited details |
| Hoxha 2020 | Quantitative descriptive | Y | Y |  |  |  |  |  | Y | Y | Y | CT | Y |  |
| Iritani 2020 | Non-randomised | Y | Y | N | CT | Y | Y | CT |  |  |  |  |  |  |
| Kennelly (2020) + | Quantitative descriptive | Y | Y |  |  |  |  |  | CT | Y | Y | N | Y |  |
| Kim (2020) | Quantitative descriptive | Y | CT |  |  |  |  |  | N | N | N | N | N |  |
| Kimball (2020) | Quantitative descriptive | Y | Y |  |  |  |  |  | Y | Y | Y | Y | Y* | *Data limited to descriptive statistics (counts/ percentages) brief report |
| Klein (2020) | Quantitative descriptive | N | N |  |  |  |  |  |  |  |  |  |  | Autopsy reporting |
| Lennon (2020) + | Quantitative descriptive | Y | Y |  |  |  |  |  | Y | CT | Y | Y | Y |  |
| Louie (2020) | Quantitative descriptive | Y | Y |  |  |  |  |  | Y | Y | Y | Y | Y* | *Data limited to descriptive statistics presented in a brief report |
| McMichael (2020b) | Quantitative descriptive | Y | Y |  |  |  |  |  | Y | Y | Y | N | Y* | *Data limited to descriptive statistics |
| Office National Statistics (2020) | Quantitative descriptive | Y | Y |  |  |  |  |  | Y | CT | Y | CT | Y |  |
| Patel (2020) | Longitudinal, Descriptive quantitative | Y | Y |  |  |  |  |  | Y | Y | Y | Y | Y |  |
| Quicke (2020) + | Quantitative descriptive | Y | Y |  |  |  |  |  | CT | CT | CT | CT | Y | Limited data reported and virologic assay. |
| Quigley (2020) | Quantitative descriptive | Y | Y |  |  |  |  |  | Y | N | Y | N | Y* | *Limited descriptive data reported in a research letter |
| Roxby (2020) JAMA | Quantitative descriptive | Y | Y |  |  |  |  |  | Y | Y | Y | Y | Y* | *Descriptive data reported |
| Sacco (2020) | Quantitative descriptive | Y | Y |  |  |  |  |  | Y | Y | Y | Y | Y |  |
| Sanchez (2020) | Quantitative descriptive | Y | Y |  |  |  |  |  | CT | CT | Y | CT | Y* | *Descriptive data reported on prevalence (counts/ percentages) |
| Stall (2020) (CMAJ) | Non-randomised | Y | Y | CT | Y | Y | CT | Y |  |  |  |  |  |  |
| Stow (2020) + | Quantitative descriptive | Y | Y |  |  |  |  |  | Y | Y | Y | Y | Y |  |
| Telford (2020) | Non-randomised | Y | Y | CT | Y | Y | N | Y |  |  |  |  |  |  |
| Unruh (2020) | Quantitative descriptive | Y | Y |  |  |  |  |  | Y | CT | Y | Y | Y |  |

Y = Yes, N= No, CT= Can’t tell

+ pre published manuscript available
